# Accurate Prediction of Breast Cancer Survival through Coherent Voting Networks with Gene Expression Profiling

**DOI:** 10.1101/2020.10.28.20221671

**Authors:** Marco Pellegrini

## Abstract

We describe a novel machine learning methodology which we call *Coherent Voting Network* (CVN) and we demonstrate its usefulness by building a 5-years prognostic predictor for post-surgery breast cancer patients based on CVNs. *Coherent Voting Network* (CVN) is a supervised learning paradigm designed explicitly to uncover non-linear, combinatorial patterns in complex data, within a statistical robust framework. Breast Cancer patients after surgery may receive several types of post-surgery adjuvant therapeutic regimen (endocrine, radio- or chemo-therapy, and combinations thereof) aiming at reducing relapse and the formation of metastases, and thus favouring log term survival. We wish to predict the outcome of adjuvant therapy using just small molecular fingerprints (mRNA) of the patient’s transcriptome. Our aim is to have simultaneously high scores for PPV (positive predictive value) and NPV (negative predictive value) as these are important indices for the final clinical applications of the predictor. A Training-validate-test protocol is applied onto CVN built on patient data from the Metabric Consortium (about 2000 patients). For the testing pool of 82 lymph node positive patients we obtain PPV 0.77 and NPV 0.78 (Odds Ratio 11.50); for the pool of 61 lymph node negative patients we obtain PPV 0.68 and NPV 0.88 (Odds Ratio 16.07). Improved results are obtained on some specific sub-types of BC. For the testing pool of 16 TNBC patients we obtain PPV 1.0 and NPV 0.83 (Odds Ratio 45.00). For the testing pool of 18 HER2+ patients we obtain PPV 0.91 and NPV 1.0 (Odds Ratio 40.00). For the testing pool of 41 Luminal B patients we obtain PPV 0.75 and NPV 0.95 (Odds Ratio 60.00). Effectiveness of the selected fingerprints is confirmed also on several independent data sets (for a total of 601 patients) from the NCBI Gene Expression Omnibus (GEO).

## 1 Introduction

It is estimated that for 2020 in the US the expected number of new cases of Breast Cancer (BC) in female patients is about 276,000 (30% of all new tumor cases in female patients) and the expected number of deaths caused by Breast Cancer in female patients is about 42,000 (15% of all deaths due to tumors in female patients), thus making BC the first type of cancer for number of new cases, and the second type of cancer as cause of death^1^ in female patients. Similar rankings are observed in Europe^2^ and China^3^.

Primary cancer treatment for new cases of BC is surgery (of various types), followed by adjuvant therapies^1^. For a patient affected by breast cancer, after tumor removal, it is necessary to decide which adjuvant therapy is able to prevent the tumor relapse and the formation of metastases. To this effect a series of measurements of several parameters (clinical, histological, molecular) are collected and evaluated by experts with the help of guidelines.

Conventional clinical-pathological parameters have been used since the definition of the first cancer staging systems in 1946^4^ up to the recent St. Gallen Consensus^5^ to select patients eligible for adjuvant treatment following BC surgery, thus helping in avoidance of unnecessary cytotoxic treatments. The high social and personal cost of chemotherapy and the evidence of over-prescription with the standard methodologies^6^, fueled the search for scientific and technological advances in this area, that could impact on clinical practice.

The need for better prognosis and prediction of therapy results has lead to substantial research in alternative biomarkers based on BC molecular profiling, and novel prediction models and algorithms, that could overcome intrinsic limitations of previous approaches. In particular high-throughput sequencing technologies have been key enablers for the success of this new approach, as well as the efforts for systematic collection of molecular data.

At this moment prognostic tools based on molecular biomarkers are considered valid clinical decision support tools, complementing traditional histopathology (see e.g. the Mammaprint and Oncotype DX tests)^7^.

Prognostic molecular tests are cost-effective versus the cost of chemotherapy for patients who would not eventually benefit from it. They are considered complementary to histology-based more traditional methods (e.g. TNM staging). Van’t Veer and her co-authors^8^ describe a panel of 70 mRNA biomarkers for breast cancer predicting survival after 5 years from breast cancer surgery. This panel is the basis for the *Mammaprint* test, which after several clinical trials, has been approved by regulatory agencies in USA and Europe for clinical use.

Paik et al.^9^ proposed a panel of 16 genes (plus 5 control genes) whose expression level is the basis for computing a score that allows to classify patients into low, medium and high risk of relapse within 5 years after surgery. This panel is commercialized as *Oncotype DX* and it has been validated in the clinical trial TAILORx^10^. In published data the intermediate class, which is rather neutral for clinical decisions, covers 30% of the patients in the testing cohort. Other methods for multigene based prognosis of breast cancer are covered in a survey by Gyo"rffy et al.^11^.

In this paper we describe a new machine learning (ML) supervised classification method and we apply it to the task of producing prognostic predictions of survival at 5 years for BC patients using gene expression levels measured from the samples of the tumor surgically removed. The prediction method is conditional on the type of post-operative adjunct therapy selected for the patient. Data from a cohort of 2000 patients^2^ available through the Metabric consortium^12^ are used to train, validate and test the prognostic predictor, and they indicate competitive performances with respect to state of the art methods.

This article is organized as follows. In Section 2 we describe the main results in the application of the CVN-based prognostic predictor on Metabric data. In Section 3 we give a high level description of the CVN method, while more details are in the Supplementary Materials. In Section 4 we compare the CVN-based prognostic predictor against other state of the art ML methods using the *Autoweka* package. In Section 5 we apply the molecular fingerprints derived for Metabric to several independent cohorts of patients. In Section 6 we comment on strong and weak points of the proposed method, as well as on possible extensions.

## 2 Results

### 2.1 Therapy classes

Patients after surgery may or may not follow one of the following adjuvant therapies: chemotherapy, radiation therapy or hormone therapy (also called endocrine therapy), which are reported in Metabric annotations. There are thus 8 possible combinations of three therapies. For each therapy profile we repeat the training-validate-testing procedure to obtain 8 therapy-specific gene sub-panels and prediction performance estimates (primary stratification)^3^. Table 1 reports 5 therapy classes for which Metabric data are sufficiently numerous to estimate the statistical significance of the predicted performance indices, and the automatic hyper-parameter/feature selection optimization converges.

**Table 1.** Performance of therapy-based stratification. Results on test data with automatic hyperparameter optimization and feature (gene) selection. Therapy class labels are (RAD, CHE, HOR). *n*.*p*. = number of patients. *n*.*a*. = number of answers. 95% Confidence Interval. *lrt p-val* = p-value for the log rank test. *lh* : lookahead number. *fp* = fingerprint size.

| Therapy | yes-no-yes | no-no-yes | no-no-no | yes-no-no | yes-yes-yes |
| --- | --- | --- | --- | --- | --- |
| n.p. | 43 | 31 | 21 | 13 | 35 |
| > 5y | 17 | 14 | 7 | 8 | 8 |
| < 5y | 26 | 17 | 14 | 5 | 27 |
| n.a. | 37 | 30 | 21 | 13 | 30 |
| Sen. | 0.65 | 0.81 | 0.66 | 0.85 | 0.8 |
| Spec. | 0.92 | 0.78 | 0.8 | 0.66 | 0.84 |
| OR | 24.3 | 16.8 | 8.0 | 12.0 | 21.0 |
| OR p-val | 0.0006 | 0.002 | 0.11 | 0.1 | 0.01 |
| CI-Lo | 2.6 | 2.55 | 0.96 | 0.79 | 1.8 |
| CI-Hi | 221 | 111 | 66 | 180 | 240 |
| kappa | 0.52 | 0.58 | 0.44 | 0.53 | 0.51 |
| AUC | 0.85 | 0.87 | 0.77 | 0.77 | 0.63 |
| AUC p-val | 0.0001 | 0.0002 | 0.02 | 0.06 | 0.13 |
| lrt p-val | 0.02 | 0.0006 | 0.06 | 0.33 | 0.03 |
| lh | 2 | 2 | 4 | 1 | 3 |
| fp | 7 | 12 | 17 | 8 | 5 |

The number of genes in each fingerprint for the therapy classes ranges from a minimum of 5 to a maximum of 17, with an average of 9.875. Overall 78 distinct genes are used. The selected fingerprints hardy overlap with previously known fingerprints (see Supplementary materials)

### 2.2 Secondary stratifications

Starting from the 5 sub-panels based on the therapy-classes (primary stratification), it is possible to define stratifications based on different features (secondary stratification) of the patient. We take into consideration ER status as measured by IHC (Table 2), Intrinsic Type (Table 3), ER/HER2 classification (Table 4), Tumor stage (Table 5), Tumor grade (Table 6), and Lymph node state (Table 7). Kaplan-Meier plots for three interesting subclasses are shown in Figures 1, 2, 3, 4, and 5. Kaplan-Meier plots for all the secondary stratifications are shown in the Supplementary materials. The secondary stratifications do not change the prediction of any single patient but provide a different evaluation of the quality of the prediction.

**Table 2.** Secondary stratification by ER status

| type | n.p. | > 5y | < 5y | n.a. | Sen. | Spe. | or | p-val | CI-Lo | CI-Hi | kappa | lrt pval | PPV | NPV |
| --- | --- | --- | --- | --- | --- | --- | --- | --- | --- | --- | --- | --- | --- | --- |
| pos | 116 | 78 | 38 | 107 | 0.67 | 0.83 | 9.83 | 6.67e-07 | 3.88 | 24.93 | 0.50 | 0.001 | 0.67 | 0.83 |
| neg | 24 | 10 | 14 | 21 | 0.86 | 0.71 | 15.00 | 0.02 | 1.63 | 138.16 | 0.57 | 0.01 | 0.86 | 0.71 |

**Table 3.** Secondary stratification by intrinsic status

| type | n.p. | > 5y | < 5y | n.a. | Sen. | Spe. | or | p-val | CI-Lo | CI-Hi | kappa | lrt pval | PPV | NPV |
| --- | --- | --- | --- | --- | --- | --- | --- | --- | --- | --- | --- | --- | --- | --- |
| LumA | 45 | 37 | 8 | 41 | 0.25 | 0.88 | 2.42 | 0.58 | 0.36 | 16.34 | 0.14 | 0.11 | 0.33 | 0.83 |
| LumB | 41 | 26 | 15 | 37 | 0.92 | 0.83 | 60.00 | 1.09e-05 | 5.98 | 601.61 | 0.72 | 0.05 | 0.75 | 0.95 |
| claudin-low | 14 | 7 | 7 | 13 | 0.71 | 0.83 | 12.50 | 0.10 | 0.84 | 186.31 | 0.54 | 0.24 | 0.83 | 0.71 |
| Her2 | 22 | 12 | 10 | 20 | 0.90 | 0.60 | 13.50 | 0.06 | 1.20 | 152.22 | 0.50 | 0.79 | 0.69 | 0.86 |
| Basal | 14 | 3 | 11 | 14 | 0.82 | 0.67 | 9.00 | 0.18 | 0.52 | 155.25 | 0.43 | 0.06 | 0.90 | 0.50 |

**Table 4.** Secondary stratification by 3 genes status

| type | n.p. | > 5y | < 5y | n.a. | Sen. | Spe. | or | p-val | CI-Lo | CI-Hi | kappa | lrt pval | PPV | NPV |
| --- | --- | --- | --- | --- | --- | --- | --- | --- | --- | --- | --- | --- | --- | --- |
| HER2+ | 18 | 8 | 10 | 15 | 1.00 | 0.80 | 40.00 | 0.01 | 1.98 | 807.14 | 0.84 | 0.01 | 0.91 | 1.00 |
| ER+/HER2- | 98 | 68 | 30 | 90 | 0.57 | 0.84 | 6.93 | 1.37e-04 | 2.53 | 19.02 | 0.42 | 0.07 | 0.62 | 0.81 |
| ER-/HER2- | 16 | 6 | 10 | 15 | 0.90 | 1.00 | 45.00 | 7.62e-03 | 2.29 | 885.65 | 0.86 | 0.004 | 1.00 | 0.83 |

**Table 5.** Secondary stratification by Tumor stage

| type | n.p. | > 5y | < 5y | n.a. | Sen. | Spe. | or | p-val | CI-Lo | CI-Hi | kappa | lrt pval | PPV | NPV |
| --- | --- | --- | --- | --- | --- | --- | --- | --- | --- | --- | --- | --- | --- | --- |
| 1 | 27 | 20 | 7 | 26 | 0.71 | 0.84 | 13.33 | 0.01 | 1.71 | 103.76 | 0.53 | 0.03 | 0.62 | 0.89 |
| 2 | 68 | 43 | 25 | 61 | 0.80 | 0.78 | 14.00 | 1.66e-05 | 3.99 | 49.16 | 0.57 | 0.009 | 0.71 | 0.85 |
| 3 | 13 | 5 | 8 | 13 | 0.62 | 1.00 | 8.33 | 0.14 | 0.63 | 110.03 | 0.56 | 0.02 | 1.00 | 0.62 |

**Table 6.** Secondary stratification by Tumor grade

| type | n.p. | > 5y | < 5y | n.a. | Sen. | Spe. | or | p-val | CI-Lo | CI-Hi | kappa | lrt pval | PPV | NPV |
| --- | --- | --- | --- | --- | --- | --- | --- | --- | --- | --- | --- | --- | --- | --- |
| 2 | 54 | 39 | 15 | 45 | 0.77 | 0.84 | 18.00 | 1.75e-04 | 3.62 | 89.58 | 0.59 | 0.0006 | 0.67 | 0.90 |
| 3 | 75 | 40 | 35 | 72 | 0.74 | 0.76 | 8.99 | 4.38e-05 | 3.09 | 26.13 | 0.50 | 0.02 | 0.74 | 0.76 |

**Table 7.** Secondary stratification by lymph node status

| type | n.p. | > 5y | < 5y | n.a. | Sen. | Spe. | or | p-val | CI-Lo | CI-Hi | kappa | lrt pval | PPV | NPV |
| --- | --- | --- | --- | --- | --- | --- | --- | --- | --- | --- | --- | --- | --- | --- |
| POS | 82 | 48 | 34 | 75 | 0.70 | 0.83 | 11.50 | 4.19e-06 | 3.83 | 34.54 | 0.54 | 0.0003 | 0.77 | 0.78 |
| NEG | 61 | 41 | 20 | 56 | 0.79 | 0.81 | 16.07 | 2.41e-05 | 4.06 | 63.63 | 0.58 | 0.005 | 0.68 | 0.88 |

**Figure 1.**
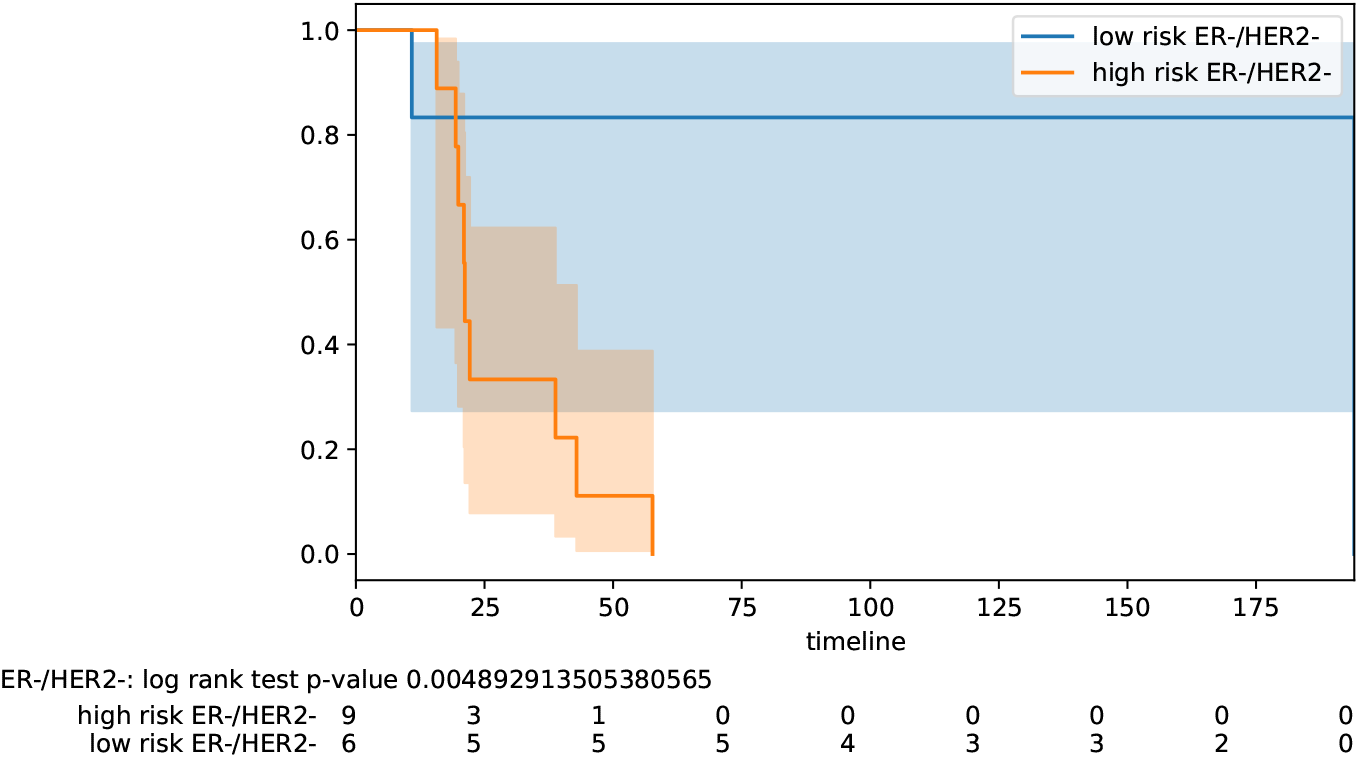
Stratification by hormonal type : ER-/Her2-.

**Figure 2.**
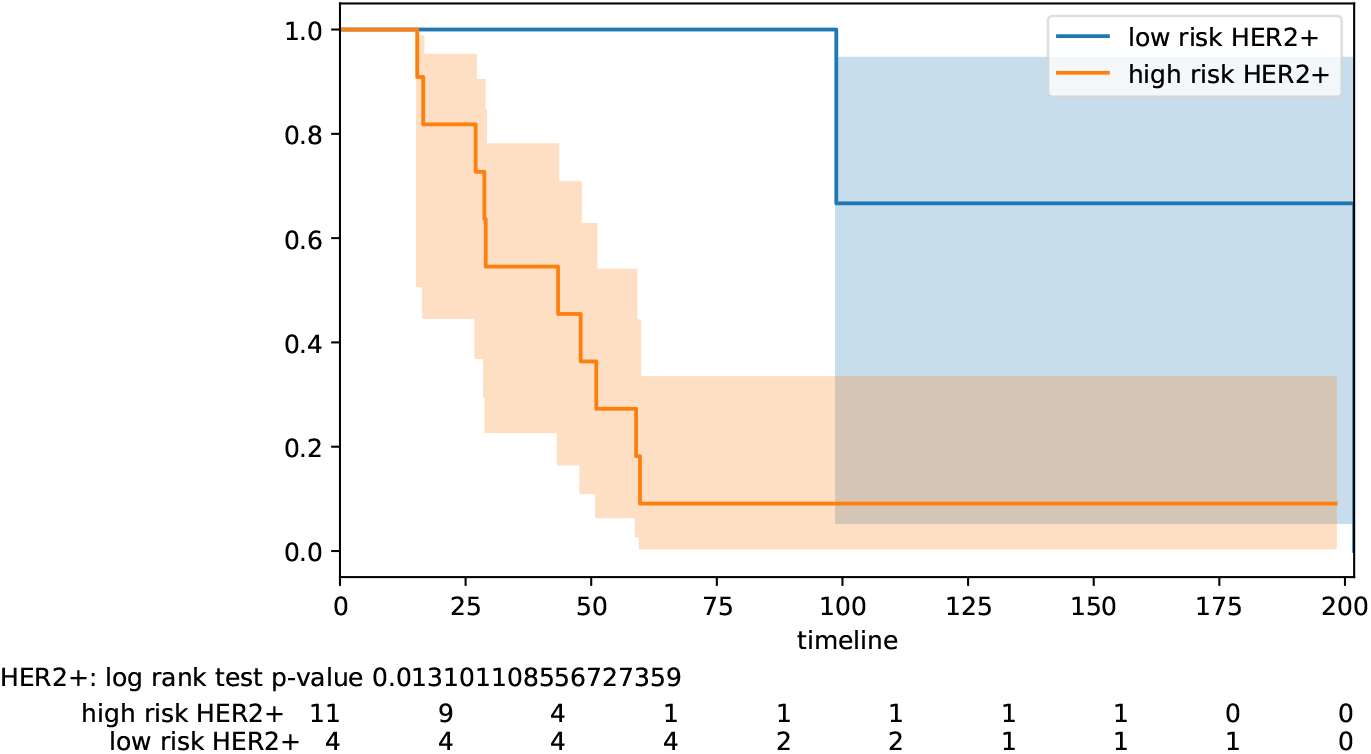
Stratification by hormonal type : Her2+.

**Figure 3.**
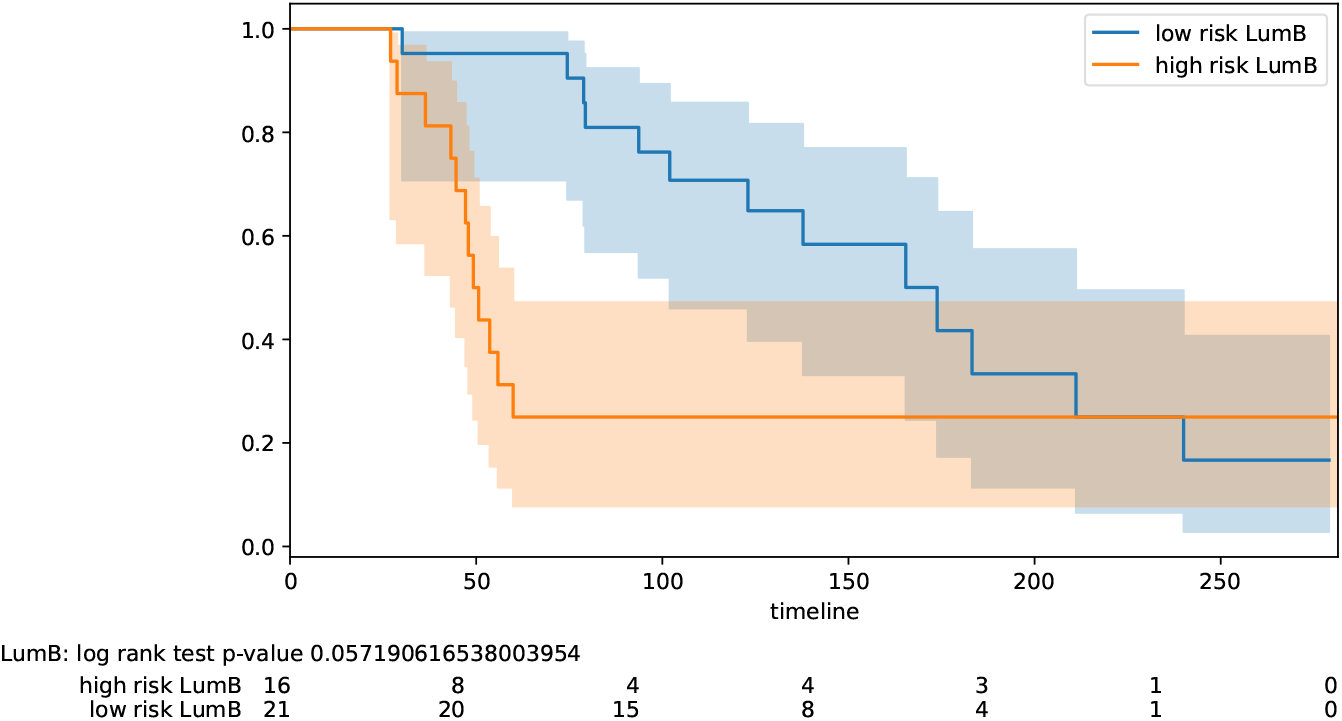
Stratification by intrinsic type : Luminal B.

**Figure 4.**
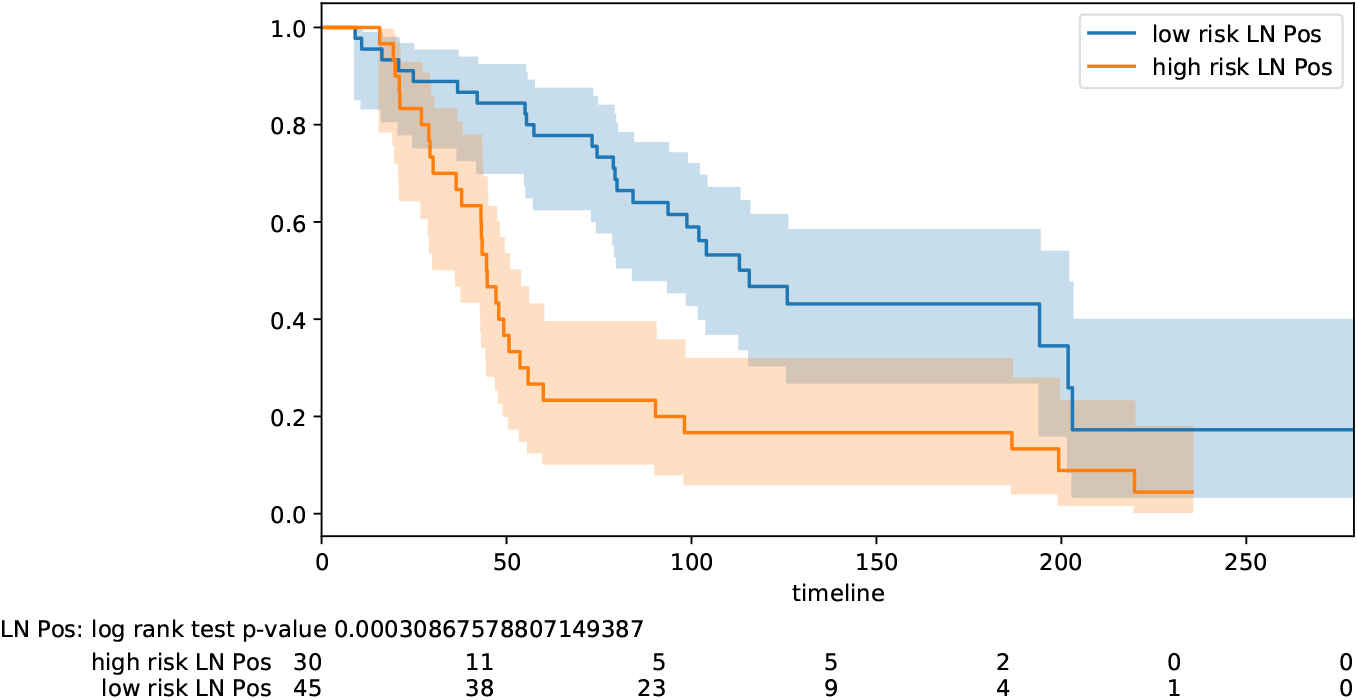
Stratification lymph node status : positive.

**Figure 5.**
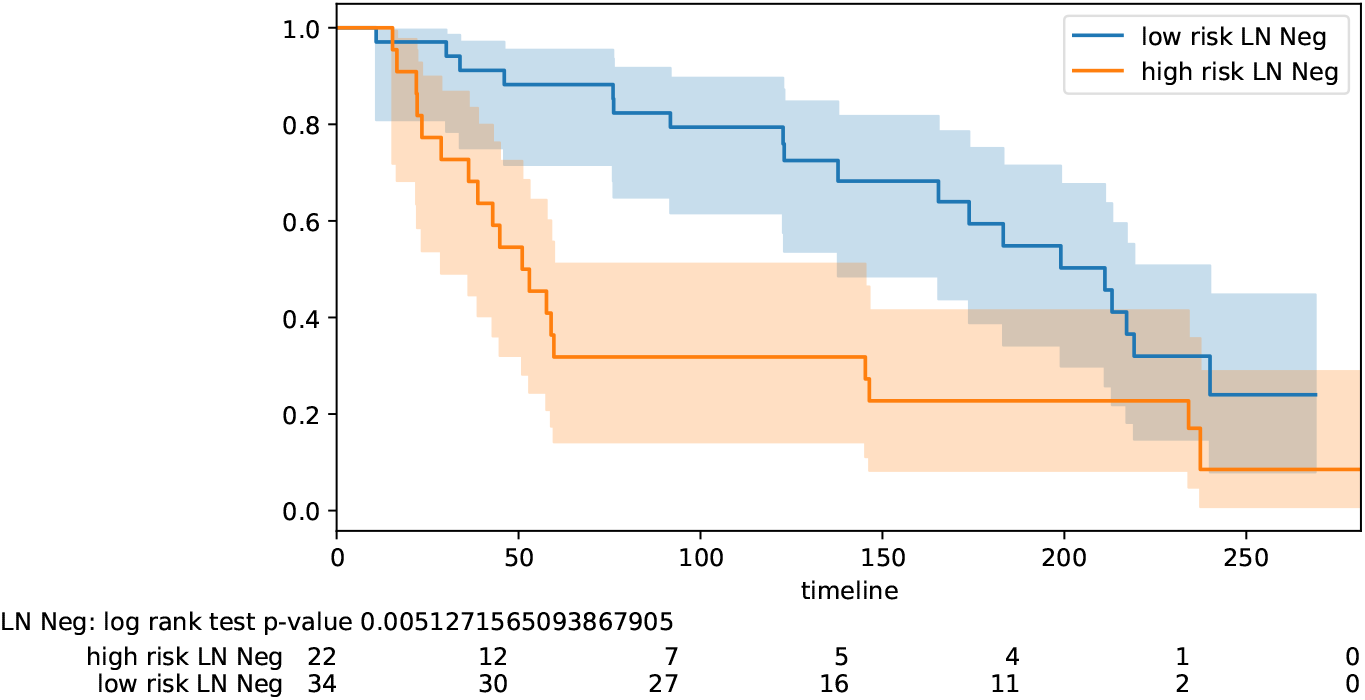
Stratification lymph node status : negative.

## 3 Methods

Here we give an overview of the Coherent Voting Network (CVN) methodology at a high level. For details we refer to the Supplementary materials (Section *Methods in detail*). The description is in two parts. The first part introduces the CVN and its use for prognosis prediction. The second part describes the feature-selection and hyper-parameter optimization procedure that is performed in a train-validate-test protocol aiming at optimizing the gene fingerprint, the CVN configuration and to estimate the performance of the method on a testing set of patients.

### 3.1 Construction of a CVN

As working with a complete gene set is a computational burden and may introduce too much noise from the experimental measure, we apply a mild initial statistical filter to preserve in the computation only genes able to discriminate the two categories of patients patients (high-risk or low-risk) that correspond to bad and good prognosis, using thresholds for fold change, t-test, ks-test (Kolmogorov-Smirnov), and mwu-test (Mann-Whitney U). Thus the gene set we use in the further CVN construction is composed of genes passing a combination of these statistical discrimination tests.

We build a bipartite graph *G* in which we have patient nodes *P* and Gene-Interval nodes *GI* where each node of the *GI* class is labelled with a gene and an interval of values for the expression of that gene. This graph is built in a straightforward manned from the input data matrix of gene expression for a pool of patients, by using quantization methods^13^. We build a partial dense cover of this bipartite graph (see definition in Pellegrini et al.^14^) which is a collection C of dense subgraphs of *G*, where each subgraph is also called a *community*. Each community will have both patients and gene node, and the communities may overlap. Let us for the moment concentrate only on the patient nodes. Each patient may belong to many communities. Each patient has a category (high-risk or low-risk) that correspond to bad and good prognosis. Each community expresses a vote (high-risk, low-risk, or null) by a voting scheme (say, for the moment, simple majority, but more schemes are described in the Supplementary Materials). Each patient receives a prediction that is the majority category expressed by the communities it belongs to. Finally the voting is coherent for a given patient *p* if the vote received by *p* is equal to her category. The degree of coherence of the voting network is the fraction of patients for which it is coherent. Ideally, the higher the degree of coherence of a CVN the better such CVN is as a basis for a predictor. The key point is that in such a construction the partial dense cover does not depend on the category of the patients, thus we may have in input non-classified patients, for which the vote of the network represents their category prediction. The intuition is that a network that is coherent for the classified patients, even if built without knowing their category, is a good predictor also for the unclassified ones.

We can see a CVN as a generalization of the notion of *guilt by association* (GbA) in biological networks. In a typical application some nodes in a biological network will have labels and some will be unlabelled. We make a prediction of an unlabelled node by using the labelled nodes with a neighborhood of the unlabelled node in the biological graph. Note that in GbA each node receives a vote from a *single subset* of the nodes.

So far each community in a CVN may have a large number of genes, and one of our aims is to find a minimal set of genes that leave the communities (of patients) unchanged since the reduction of the number of genes would not change much their density. To achieve this goal we consider now only the genes belonging to any community. We look for a minimal set *M* of genes so that each community (of genes) includes at least *k* genes in *M*. The set *M* can be well approximated by a using a greedy set multi-cover algorithm (see e.g.^15^).

After computing the minimal set *M* of genes we can rebuild the CVN using only the patient set *P* and the genes in *M* obtaining a CVN’, measure the coherence of CVN’, and use CVN’ for prediction of the category of unclassified patients.

### 3.2 Train-validate-test protocol

Each phase of the construction described above depends on the choice of values for hyper-parameters, and we will have a CVN for each such choice (which we call a parameter-vector *v* of the parameter-space *V*). While sophisticated strategies for searching this discrete parameter-space exist (in ML they are termed *hyper-parameter optimization strategies*) in our application the construction of a single CVN is in practice very efficient thus we will use *greedy search* and compute a CVN for each *v* ∈ *V*, as |*V*| is in the range of only a few hundreds.

A further aim, besides finding an optimal *v*, and a small gene set *M* is to have high performance for the testing phase in a train-validation-test set-up.

We begin by splitting the initial set of patients into three sets: the training set *T*0, the validation set *T*1 and the test set *T*2. In a standard ML setting information leaking is avoided by finding the optimal (*v*\*, *M*\*) pair only on (*T*0, *T*1) and then applying such optimal predictor to (*T*0, *T*2). The performance is measured on this unique predictor for (*T*0, *T*2). We relax such all/nothing schema by allowing the use of *T*2 in the choice of (*v*\*, *M*\*) in a very limited and controlled way, by use of the concept of *lookahead*. Instead of producing a single predictor on (*T*0, *T*2) we produce a ranking of all predictors on (*T*0, *T*1) that we can build by choosing a *v* ∈ *V*. We then lookup vectors *v* in this ranked list, and we stop when the corresponding predictor for (*T*0, *T*2) satisfy a stopping criterion. The number of vectors *v* we visit in this lookahead process is the *lookahead number* (lh). For lh=1 we have the standard ML set up. In Table 1 we report the lh values observed for the therapy classes: 1 once, 2 twice, 3 once and 4 once.

## 4 Comparison of CVN with other ML classification methods

In order to compare our algorithmic solution with the state-of-the-art in machine learning we performed experiments with the Autoweka package (^16^,^17^) within the Weka workbench environment (^18^,^19^). Autoweka performs automatically feature selection and hyper-parameter optimization of 27 base classification methods, 10 meta-methods, and two ensemble methods, moreover it uses several feature selection search methods along with 8 feature evaluation functions. The hyper-parameters are optimized in Autoweka using a Bayesian optimization strategy to explore the space of parameters. As we noticed that the initial feature selection phase is onerous when applied to the input of roughly 24,000 genes, we also applied explicitly several Weka feature selection pre-filters so to reduce the number of features in input to Autoweka. Autoweka uses ten-fold cross-validation over the training set to select the best configuration of hyper-parameters. We fixed the kappa statistics as the objective function to be maximized in the learning phase (see Supplementary Materials). The reported kappa statistics is computed on the trained predictor for the test data set.

Table 8 reports the kappa statistics for the best Autoweka trained classifiers (in round brackets) along with the result we obtain with the coherent voting networks over the test set. For CVN next to the kappa statistics we report the lookahead number or, in two cases, the manual selection of the best configuration. Ignoring for the moment the two manually selected configurations, we notice that we can get the highest kappa values in three of the six remaining cases. We also notice for CVN a robust uniform behaviour with consistent high positive values of kappa. In all columns (except corr-ranker) there are negative entries indicating that the best ML method for that input has performance worse than a random classifier. For the *corr-ranker* feature selection the ML methods have all positive values, but generally lower than those of CVN. Moreover, the best Autoweka results are attained by 15 different methods thus making it hard to pinpoint a single winner algorithm in the Autoweka suite.

**Table 8.** Kappa statistics for training data sets for various Autoweka/Weka feature selection settings. Therapy class (RAD, CHE, HOR). *lh* : lookahead number or manually determined (m). Legend for autoweka methods: rf = Random Forest, mp = Multilevel Perceptron, nb = Naive Bayes, bn = Bayes Net, sgd = Stochastic Gradient Descent, rc = Random Committee, ibk = k-Nearest Neighbour Classifier, sl = Simple Logistic, nbm = Naive Bayes Multinomial, rpt = Fast Decision Tree REPTree (C4.5), smo = Fast Training Support Vector Machine, lo = Logistic, lwl = Locally Weighted Learning, ab = AdaBoostM1, rss = Random Sub Space. dt = Decision Table. (*) result for the validation dataset.

| Therapy | No filter | cfs-best | cfs-greedy | corr-ranker | gain-ranker | j48-ranker | j48-greedy | CVN (lh) |
| --- | --- | --- | --- | --- | --- | --- | --- | --- |
| yesnoyes | 0.30 (rf) | 0.52 (mp) | 0.52 (mp) | 0.23 (lo) | 0.38 (smo) | 0.33 (lwl) | **0.58** (mp) | 0.52 (2) |
| nonoyes | 0.35 (sl) | 0.34 (nb) | 0.15 (rf) | 0.22 (smo) | 0.16 (lwl) | 0.35 (bn) | 0.15 (rf) | **0.58** (2) |
| nonono | 0.09 (rf) | **0.64** (bn) | 0.5 (smo) | 0.35 (ibk) | 0.35 (nb) | 0.35 (nb) | 0.50 (rf) | 0.44 (4) |
| yesnono | **0.7** (rf) | 0.53 (sgd) | 0.69 (rf) | 0.39 (lo) | 0.56 (rf) | 0.56 (rf) | **0.7** (rf) | 0.53 (1) |
| yesyesno | -0.09 (dt) | 0.36 (rc) | 0.05 (nb) | 0.22 (lwl) | 0.10 (lwl) | 0.0 (ab) | 0.05 (nb) | **0.48** (m) |
| yesyesyes | -0.07 (nbm) | -0.07 (ibk) | -0.01 (ibk) | 0.19 (smo) | 0.14 (lwl) | 0.26 (rss) | -0.07 (ibk) | **0.51** (3) |
| noyesno | -0.26 (rf) | -0.03 (mp) | -0.03 (mp) | 0.11 (mp) | -0.26 (smo) | -0.22 (mp) | -0.03 (mp) | **0.41** (2) |
| noyesyes(*) | 0.0 (rpt) | -0.53 (rf) | -0.53 (rf) | 0.13 (rf) | 0.17 (rf) | 0.23 (rf) | -0.54 (rf) | **0.60** (m) |

Overall the setup experimental conditions for the Autoweka and CVN differ in some aspects, therefore the findings must be considered with care. Keeping these differences in mind, we can conclude that CVN has a level of performance at least comparable with existing ML methods methods. Moreover CVN is a single easy-to-explain method that allows for a more uniform approach to the BC prognosis problem over a wide spectrum of clinical conditions.

## 5 Performance of CVN on independent cohorts of patients

After screening of the breast cancer data sets in the NCBI GEO (Gene Expression Omnibus) repository we have identified a few BC data sets with characteristics compatible with the Metabric data set regarding the recorded therapy, end point survival (preferentially overall survival). The prediction performance is tested in a leave-one-out evaluation framework in which the multi-gene fingerprint is the optimal fingerprint defined on Metabric data. Greedy hyper-parameter optimization is applied and the best result in terms of OR subject to slackness below 15% is the selected configuration reported in the Table 9. Due to different microarray technologies, we have mapped the genes onto the probes for the target technology (using all mapping probes, if multiple probes map onto the same HUGO gene ID).

**Table 9.** Independent cohorts. Results of leave-one-out evaluation with optimal multigene fingerprints derived from Metabric data sets. Therapy class : (RAD, CHE, HOR). End point (e.p.) is os= overall survival„ dfs=disease free survival, rfs=relapse free survival. Confidende Interval for odds ratio at 95% confidence interval. n.a. = number of answers.

| GEO | 45255(ch) | 45255(ho) | 45255(chho) | 37181 | 7390 | 2034 |
| --- | --- | --- | --- | --- | --- | --- |
| end point | os | os | os | dfs | os | rfs |
| therapy | no-yes-no | no-no-yes | no-yes-yes | no-no-no | no-no-no | yes-no-no |
| n.p. | 8 | 16 | 13 | 119 | 181 | 264 |
| > 5y | 3 | 6 | 4 | 59 | 24 | 95 |
| < 5y | 5 | 10 | 9 | 60 | 157 | 169 |
| n.a. | 8 | 16 | 13 | 106 | 179 | 258 |
| kappa | 1.0 | 0.58 | 1.0 | 0.35 | 0.36 | 0.12 |
| Sen. | 1.0 | 0.8 | 1.0 | 0.66 | 0.7 | 0.90 |
| Spe. | 1.0 | 0.81 | 1.0 | 0.70 | 0.89 | 0.67 |
| OR | 15.0 | 18.0 | 36.0 | 4.59 | 20.86 | 20.12 |
| CI-Lo | 0.66 | 1.24 | 1.77 | 2.01 | 4.93 | 2.53 |
| CI-Hi | 339 | 260 | 731 | 10 | 88.2 | 159 |
| OR p-val | 0.19 | 0.03 | 0.01 | 3.7E-4 | 3.0E-5 | 1.8E-4 |
| AUC | 1.0 | 0.89 | 1.0 | 0.72 | 0.70 | 0.63 |
| AUC p-val | 0.01 | 0.005 | 0.003 | 2.6E-5 | 7.5E-4 | 1.0E-4 |

The GSE45255 data set holds information on three different therapy classes. The numbers of patients in an each class are however rather small. While for the GEO45255 chemotherapy (ch) subset there is perfect performance, the p-value for OR is too high for claiming statistical significance on this measure, but, in contrast, the AUC value is statistical significant. For the GEO45255 endocrine (ho) and the GEO45255 endocrine plus chemotherapy (chho) subsets we attain high values in kappa and OR, with a significant OR and AUC p-values.

Data set GSE37181 holds a large number of patients (119), and it is perfectly balanced among the two classes (60 vs 59), but the end point is disease free survival (dfs), rather than overall survival (os). We notice a loss in terms of OR although the kappa statistics are AUC are still in an acceptable range, with good statistical significance.

Data set GSE7390 holds a larger number of patients (181), but is unbalanced among the two classes (157 vs 24). This has the effect of a relatively low kappa statistics, however the odds ratio (20.86), sensitivity (0.7), specificity (0.89) and the p-values indicate a good performance on these indices.

Data set GSE2034 is the largest independent cohort (264) in this table and is roughly balanced (95 vs 169) within a factor.

2. Although the kappa statistics is low, the odds ratio OR is high (20.12) even if the end point is relapse free survival (rfs) rather than overall survival (os).

Overall these experiment show that the selected multi-gene fingerprints may be effective across different microarray platform and different patient cohorts, while some loss of performance can be expected when a different endpoint is used. This suggests that when we change the end point of the prediction (e.g. disease-free survival) we should recalibrate the fingerprints in the chosen setting.

## 6 Discussion

We have developed a new ML supervised classification method called *Coherent Voting Networks* (CVN) which is suitable for handling highly non-linear phenomena such as those prevalent in biological systems. We have applied CVN to the problem of predicting prognosis of BC patients in dependence of the chosen post-surgery adjuvant therapy selected. After surgery a breast cancer patient must follow a therapeutic regime aimed at preventing relapse and formation of metastases. The CVN-based prognostic tool is able to predict, with good accuracy for a large percentage of the patients, whether the patient will survive more or less than 5 years following current the state of the art adjuvant therapeutic protocols (based on chemotherapy, radiation therapy and hormone therapy). Such prognostic tool helps the clinician and the patient by validating the chosen therapeutic path (in case of predicted good prognosis), or by suggesting, in combination with other elements, the need for further investigations, or the application of newer, possibly experimental, alternative protocols (in case of predicted poor prognosis). The advantage for the patient is the possibility to personalize the therapeutic choices by using her molecular prognostic profile, with higher chance of an effective cure and survival. The advantage for the clinician is a tool to validate base-line therapeutic choices (or suggest the need for alternatives). The advantage for the health system at large, is a better discrimination among those patients requiring expensive and invasive cures (e.g. chemotherapy), and those that would benefit from less expensive and invasive ones (e.g. hormonal therapy). The CVN-based prognostic tool uses a small molecular profile of a few dozen genes that can be measured for each patient’s tumor biopsy with standard technologies like RNA-seq or RT-PCR.

The fingerprint gene panel has been identified using public data of the project Metabric (Molecular Taxonomy of Breast Cancer International Consortium) and and tested using other publicly available data of independent cohorts. Thus the results in this paper rely rather heavily on the quality of the Metabric protocols for collecting molecular and clinical data. An interesting line of research to be developed is to assess the robustness of the CVN-based prediction when different technologies and different data processing protocols are used. Preliminary tests on independent cohorts (see Table 9) suggest that the devised gene fingerprint is rather robust w.r.t changes in the gene expression measurement technology and are even capable of operating with end points different from the default one chosen in this study (overall survival). However the hyper-parameter optimization phase during predictor’s training is likely to be rather more data and technology dependent, and thus probably adoption of different technology/protocol in data collection may entail a re-training of the predictor. A second limitation of the method in it training phase is that it relies on knowledge of the adjuvant therapy chosen for the patients, with the assumption that over the time-frame of the data collection no drastic changes in the clinical practice and criteria would take place. As this cannot be guaranteed over a long period (and indeed changing current clinical protocols is the final aim of this tool) there is the practical need of a continuous monitoring to ensure consistency between the patient population used in training and the population for which the tool is applied.

The CVN methodology is a general ML supervised classification tool, and, for prognostic purposes, it can be in principle applied to many variants of this problem.

CVN-based prognostic tool is currently optimized to maximize and balance the kappa statistics (alternatively the odds ratio) across training, validation and test data, while limiting the number of patients for which no answer is given. This strategy produces also often a balancing of PPV and NPV. It is possible to obtain alternative gene panels for specific situation (or different predictors on the same panels) that may optimize directly PPV and NPV, say by maximizing PPV subject to a lower bound on NPV (or viceversa).

Also, when a higher rate of no-answers is allowed we can increase the PPV and NPV of for the given answers. Preliminary data for certain therapy classes give an NPV and PPV close to 95% for 50% of the patients. Thus with the same data it is possible to devise a cascade of predictors having higher guarantees for the easier cases, so to cover a given population by several stratified predictors (from the easiest to the most complex cases to predict).

It is possible in principle to apply this CVN methodology to derive a prognostic panel at 10 years (this information has also clinical relevance in long term follow ups).

In general it should be possible to derive similar gene panels for other tumors, provided that Metabric-like high quality data is available on a sufficiently large cohort of patients.

Finally, since we have used only gene expression data (and knowledge on the patients 5-year survival) to build the predictors, one may think that feeding other clinical or molecular indices as additional input to the CVN may improve the predictive powers. Preliminary experiments in this direction however show that a straightforward integration of known single clinical measurements do not improve predictions significantly. It remains thus open the question whether more sophisticated heterogeneous data integration strategies taking several indices at once may be beneficial within the CVN approach to prognosis predictions. A promising line of future research involves integrating mRNA and miRNA to produce mixed prognostic signatures^20,21^. Data on miRNA expression in Metabric patient’s samples has been produced recently within the *Metabric miRNA landscape project*^4^. Preliminary results from this projet indicate that “breast cancer miRNAs appear to act as modulators of mRNA-mRNA interactions rather than molecular switches". Thus while it is likely that mixed miRNA-mRNA fingerprints may sharpen some of our results, within the CVN framework, we expect that mRNA will continue to be key elements of the predictors, even in this extended setting. Certainly a better appreciation of miRNA-mRNA interactions in BC may shed more light in the causative elements of BC progression. A second promising direction of research integrates biomedical imaging and molecular profiling for prognostic purposes^22,23^.

Triple-negative breast cancer (TNBC) is an aggressive type of breast cancer affecting about 15% of the cases, and it is known to be quite non-homogeneous from a clinical and molecular point of view^24–26^. Research on devising prognostic molecular fingerprints for TNBC has thus been directed mainly at subclasses of of TNBC^27–33^. In our results we are able to attain good performance in terms of PPV, NPV and OR on the full pool of Metabric TNBC patients. The good overall performance may be explained with the intuition that the initial therapy-based stratification of the patients is able to capture implicitly the TNBC molecular and clinical heterogeneity.

HER2 positive BC covers about 25% of the BC cases. It is considered an aggressive tumoral form, and while it responds well to recent therapeutics, it is known to develop drug resistance in time and in about 50% of the cases distant metastases occur^34–37^. Molecular signatures for HER2-positive BC prognosis have been found for certain subtypes of the disease or for predicting the response to specific drugs^38–41^. Also for this important type of BC we could attain high PPV, NPV and OR results.

Lumina-B BC is one of the intrinsic types of BC discovered by Perou et al.^42^, based on clustering of BC gene expression profiles. Prognostic properties of this subtye have been investigated in particular with respect to the other intrinsic types^43^,^44^. In general however less is known about discriminating prognosis within the type^45,46^. Here we show that the CVN-based classifier is effective in discriminating good and poor prognosis patients with high PPV, NVP and OR. van de Vijver et al.^47^ report the performance of a 70-genes prognostic gene fingerprint: for lymph node negative OR patients OR is 15.0 (3.3 - 56, pval *<* 0.001) with PPV 0.63 and NPV 0.89. For lymph node positive patients OR is 13.7 (3.1 - 61, pval *<* 0.001), with PPV = 0.4 and NPV 0.95. Overall out results for lymph node positive and negative are similar in terms of OR, but in our case we have a better balancing between the PPV and NPV measures.

Paik et al.^9^ developed a 21-gene signature (16 predictive and 5 control genes) to predict recurrence in node-negative breast cancer treated with Tamoxifen, which was later incorporated in the Oncotype DX prognostic kit. Taking into account only the low and high risk classification of the patients we obtain an OR = 5.67 (3.39 - 9.46, pval 9.6e-12) with NPV 0.90 and PPV 0.38. Again, our result show a better balancing of PPV and NPV values.

Our work has focussed on selecting relatively small fingerprints that can be used to build predictive CVN, by maximizing the kappa statistic (or the odds ratio) in testing sets of patient data, subject to an upper bound on the slackness of the method (percentage of no responses). In this research we did not aim at uncovering *causative* fingerprints (i.e. a pattern of gene expression level measures that *explain* the future survival in combination with a therapeutic regime^48^. Although we cannot rule out that the uncovered genes may indeed be involved in the causation of the disease, two orders of considerations advise caution. One consideration is that a number of just slightly sub-optimal fingerprints may also be found (a phenomenon compatible also with the findings by Venet et al.^49^). Thus causative genes may be present outside a predictive fingerprint of minimal size, with an explanatory role as important as that of those present in the fingerprint. The second consideration, is that we have used one mRNA data set from protein coding genes as our feature space. It is known that BC involves several layers of biological regulation (e.g. genetic aberrations, actions of non coding RNA, epigenetic signals, multi-cell signalling, metabolic and environmental conditions), thus a causative explanation might involve a more complex interplay of several layers. Finally we did not touch yet on the topic of whether such fingerprints contain directly actionable targets for therapeutic agents (either for those drugs actually administered, or for new drugs in relation to the personal molecular profile of the patients). These related problems are of interest and may entail the collection and fusion of additional relevant ‘omic’ data, as well as the refinement of the algorithms introduced in this study.

## Data Availability

Data is available at https://github.com/MarcoPellegriniCNR/Coherent-Voting-Network-for-BC-prognosis

https://github.com/MarcoPellegriniCNR/Coherent-Voting-Network-for-BC-prognosis

## Footnotes

1 https://www.gov.uk/government/publications/chemotherapy-radiotherapy-and-surgical-tumour-resections-in-england/chemotherapy-radiotherapy-and-surgical-tumour-resections-in-england

2 See Supplementary materials for basic statistics of the main features of the populations used for training, validation, and testing.

3 See Supplementary materials for a self-contained recollection of the performance measures used in this context.

4 https://ega-archive.org/studies/EGAS00000000122

