## Supplementary-Materials for "Accurate Prediction of Breast Cancer Survival through Coherent Voting Networks with Gene Expression Profiling"

M. Pellegrini*

October 28, 2020

**2 Overlap of our fingerprints with previously published multi-gene fingerprints 16**

**3 Statistics on patient sets 17**

**4 Features of independent cohorts 22**

**5 Measures of performance 23**

*Istituto di Informatica e Telematica del CNR, Via G. Moruzzi 1, 56100-Pisa (Italy).

**6 Method in detail: Construction of Coherent Voting Networks** **25**

**7 Extending the repertoire of voting schemes** **28**

**8 Method in detail: Training and optimization of Coherent Voting Networks** **29**

**9 Manual fingerprint and hyperparameter optimization** **30**

**10 Avoiding overfitting** **31**

**11 Data, Software and tools used** **32**

### 1 Kaplan–Meier plots of therapy-based and secondary stratifications

#### 1.1 Therapy-based stratification

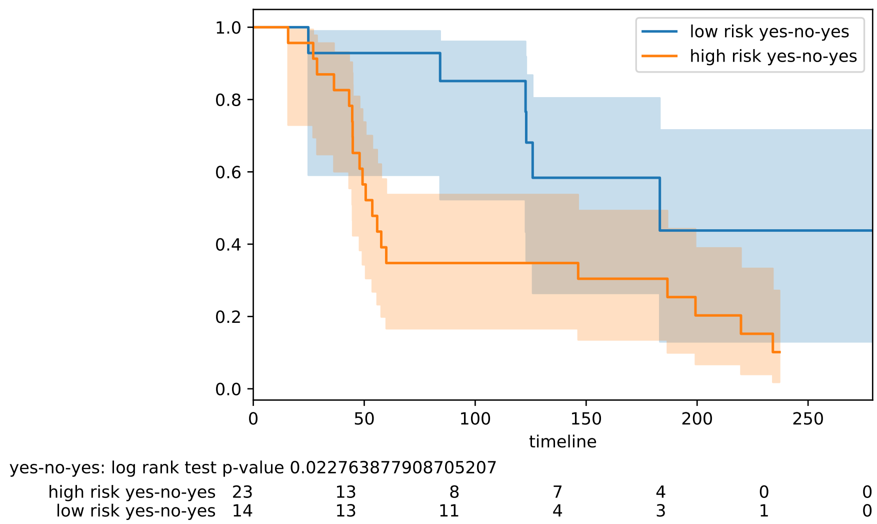

Figure 1: Stratification by therapy class (RAD, CHE, HOR) yes-no-yes.

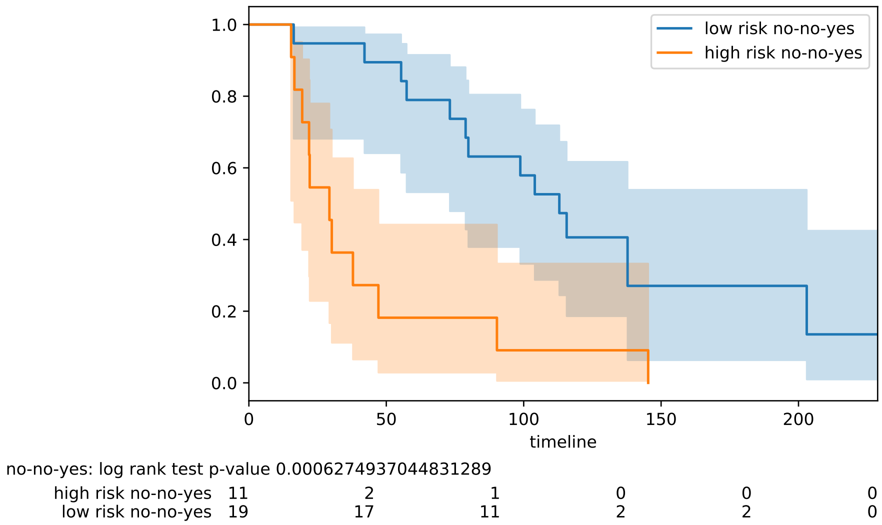

Figure 2: Stratification by therapy class (RAD, CHE, HOR) no-no-yes.

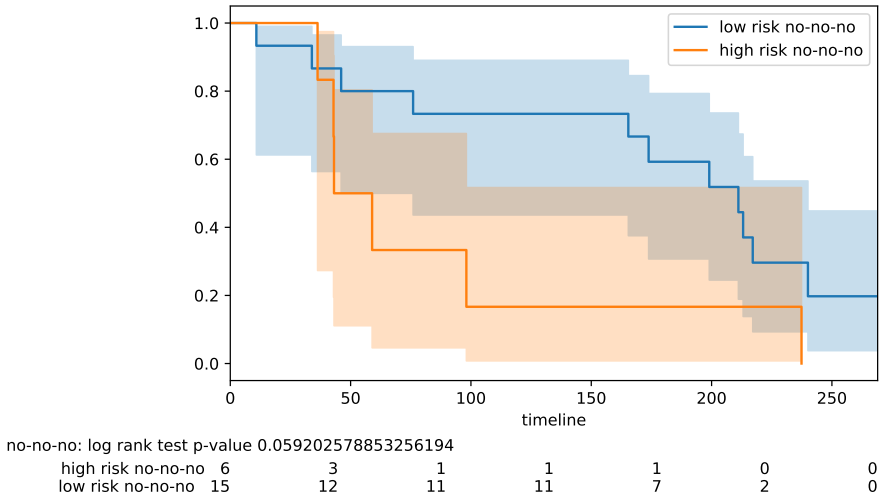

Figure 3: Stratification by therapy class (RAD, CHE, HOR) no-no-no.

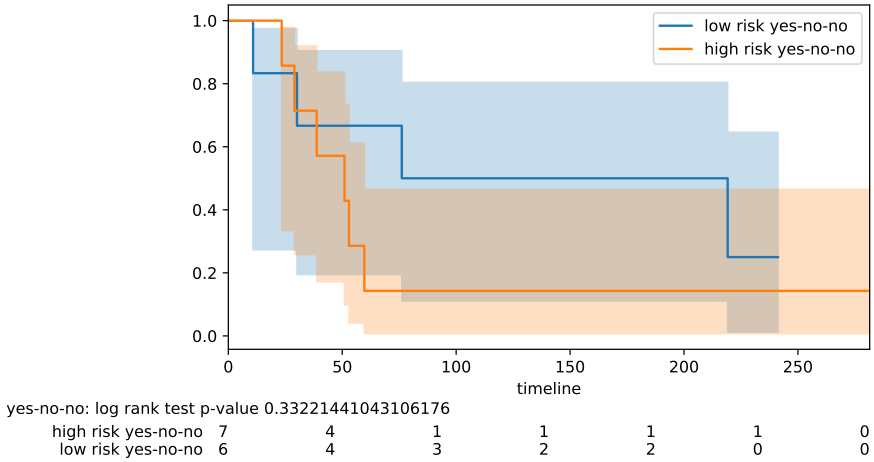

Figure 4: Stratification by therapy class (RAD, CHE, HOR) yes-no-no.

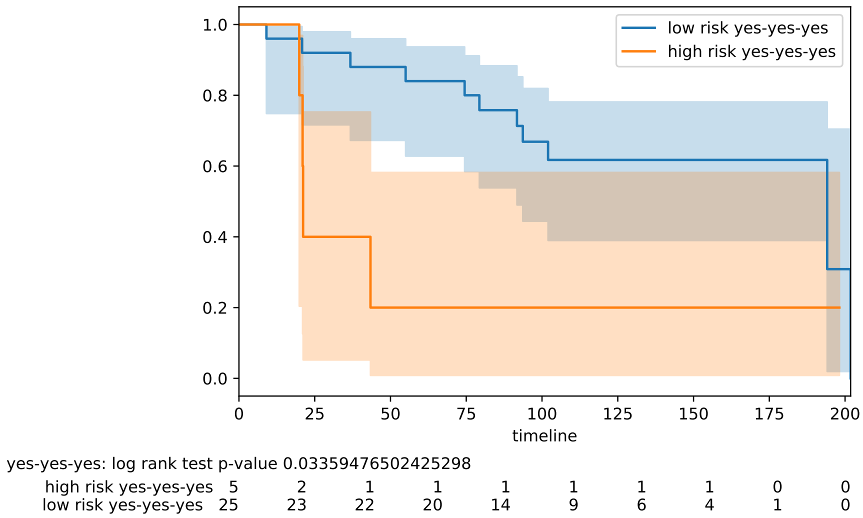

Figure 5: Stratification by therapy class (RAD, CHE, HOR) yes-yes-yes.

#### 1.2 Secondary stratifications

##### 1.2.1 Intrinsic type

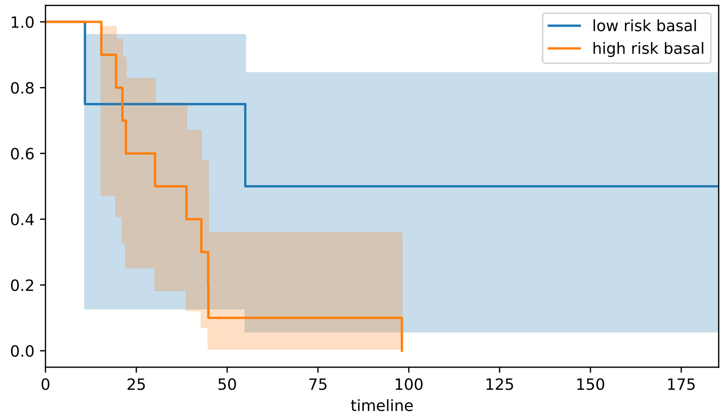

basal: log rank test p-value 0.06236264616950541

| | | | | | | | | | |
|---|---|---|---|---|---|---|---|---|---|
| high risk basal | 10 | 6 | 1 | 1 | 0 | 0 | 0 | 0 | 0 |
| low risk basal | 4 | 3 | 3 | 2 | 2 | 2 | 2 | 1 | 0 |

Figure 6: Stratification by intrinsic type : basal.

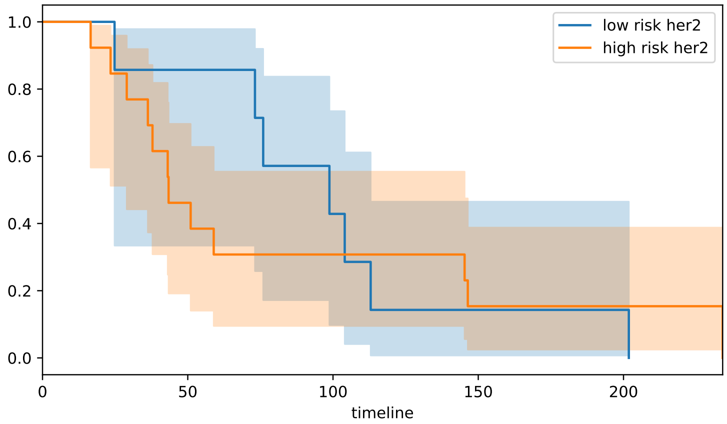

her2: log rank test p-value 0.7926616081540957

| | | | | | | |
|---|---|---|---|---|---|---|
| high risk her2 | 13 | 6 | 4 | 2 | 1 | 0 |
| low risk her2 | 7 | 6 | 3 | 1 | 1 | 0 |

Figure 7: Stratification by intrinsic type : her2.

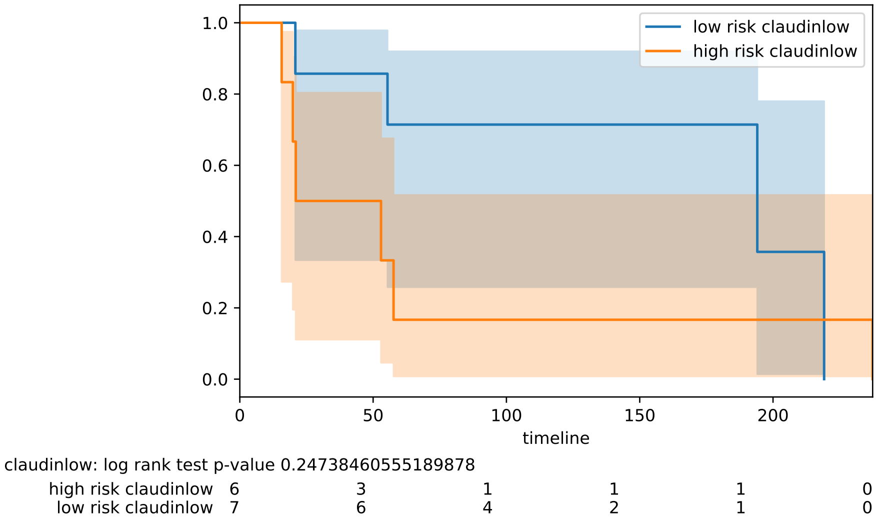

Figure 8: Stratification by intrinsic type : claudin low.

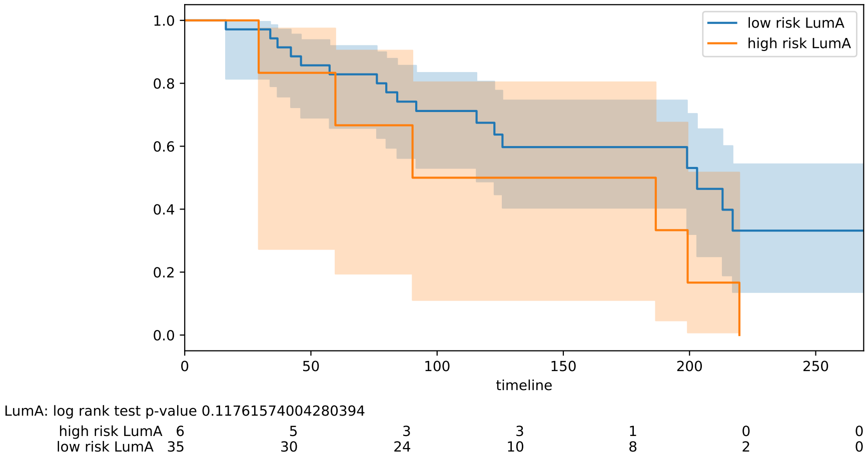

Figure 9: Stratification by intrinsic type : Luminal A.

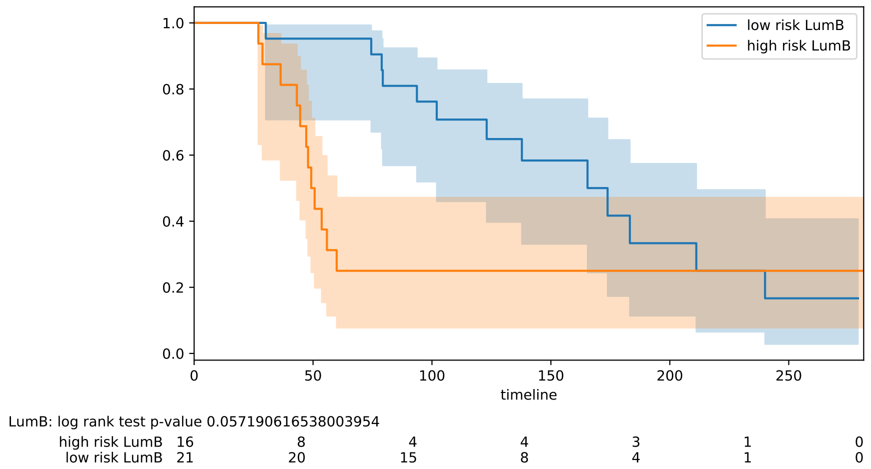

Figure 10: Stratification by intrinsic type : Luminal B.

##### 1.2.2 Hormonal type

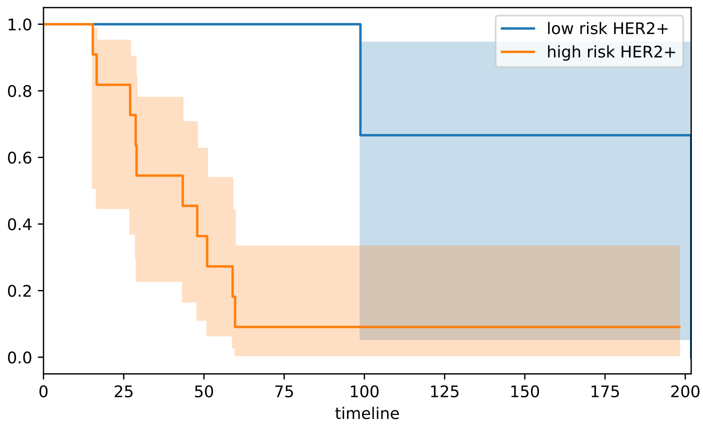

HER2+: log rank test p-value 0.013101108556727359

| | | | | | | | | | | |
|---|---|---|---|---|---|---|---|---|---|---|
| high risk HER2+ | 11 | 9 | 4 | 1 | 1 | 1 | 1 | 1 | 0 | 0 |
| low risk HER2+ | 4 | 4 | 4 | 4 | 2 | 2 | 1 | 1 | 1 | 0 |

Figure 11: Stratification by hormonal type : Her2+.

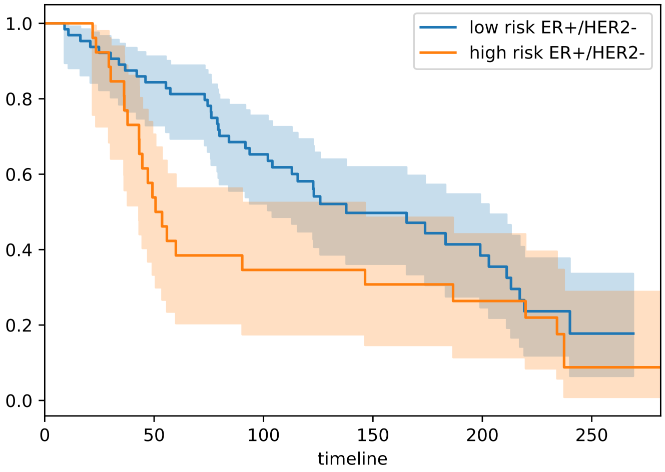

ER+/HER2-: log rank test p-value 0.07377394696561261

| | | | | | | | |
|---|---|---|---|---|---|---|---|
| high risk ER+/HER2- | 26 | 14 | 9 | 8 | 6 | 1 | 0 |
| low risk ER+/HER2- | 64 | 54 | 40 | 19 | 13 | 2 | 0 |

Figure 12: Stratification by hormonal type : ER+/Her2-.

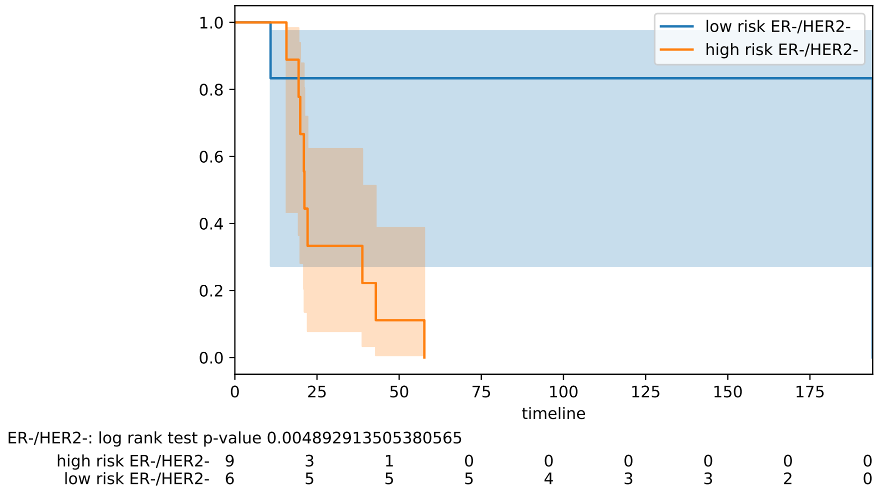

Figure 13: Stratification by hormonal type : ER-/Her2-.

##### 1.2.3 Hormonal ER by IHC

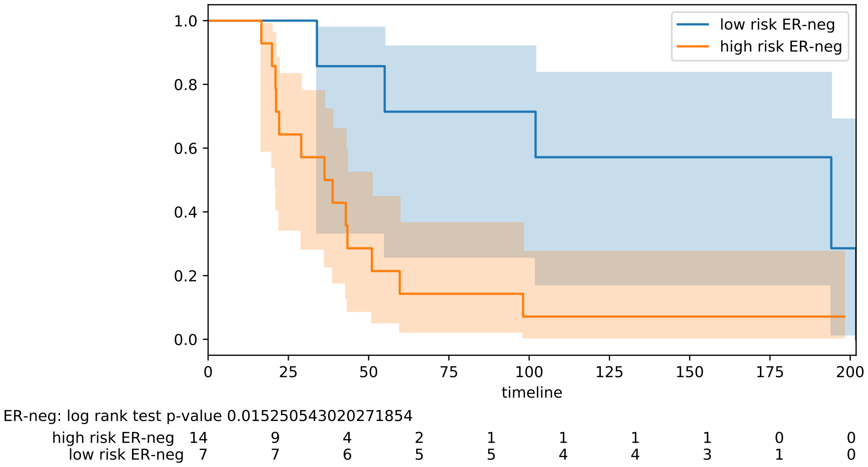

Figure 14: Stratification by IHC ER type : ER-.

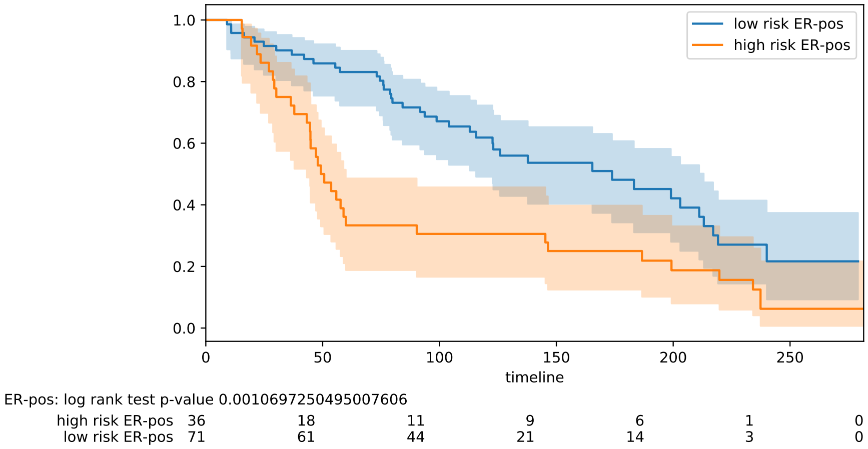

Figure 15: Stratification by IHC ER type : ER+.

##### 1.2.4 Lymph node status

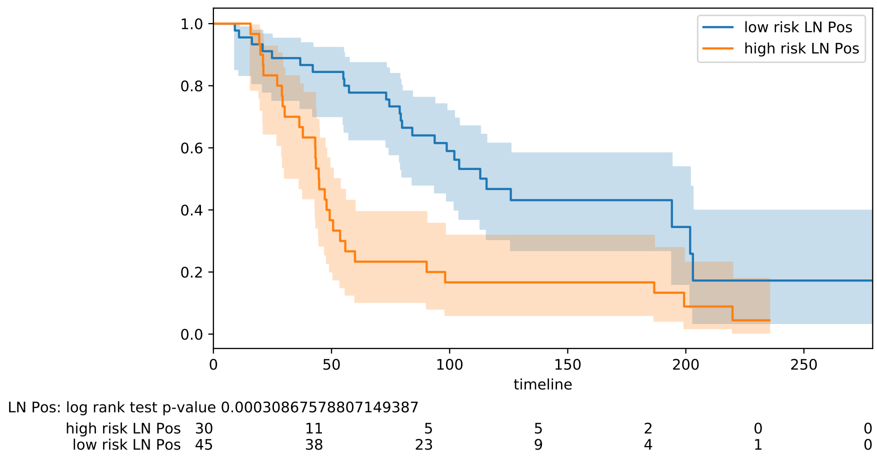

Figure 16: Stratification lymph node status : positive.

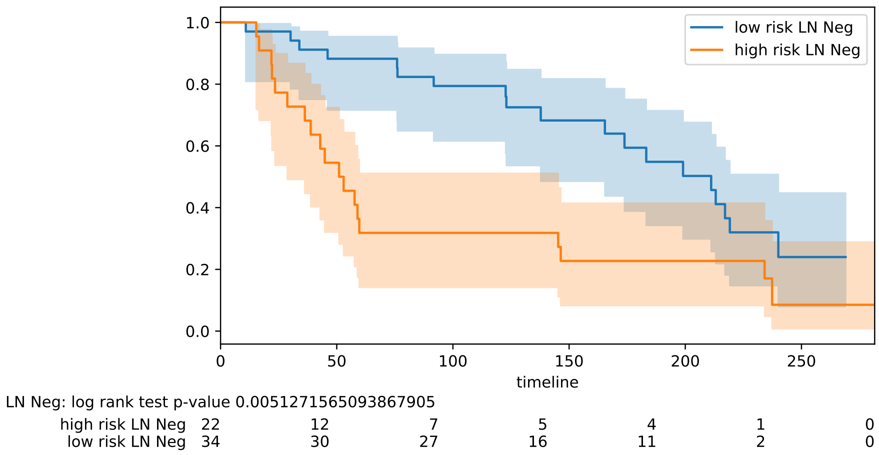

Figure 17: Stratification lymph node status : negative.

##### 1.2.5 Tumor grade status

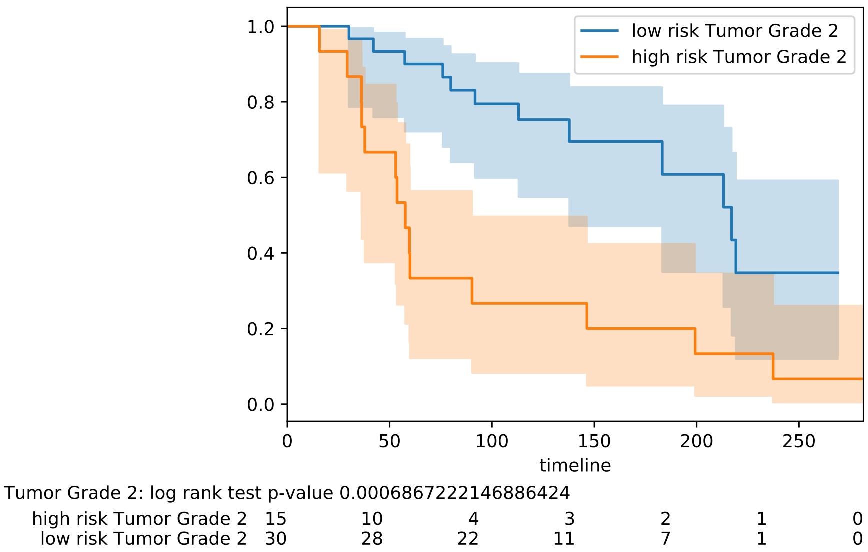

Figure 18: Stratification tumor grade : 2

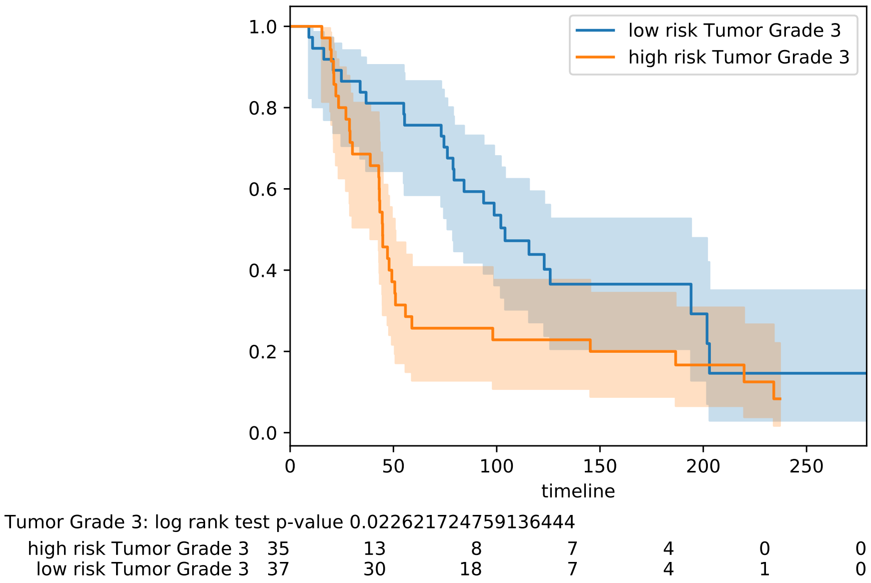

Figure 19: Stratification tumor grade : 3

##### 1.2.6 Tumor stage status

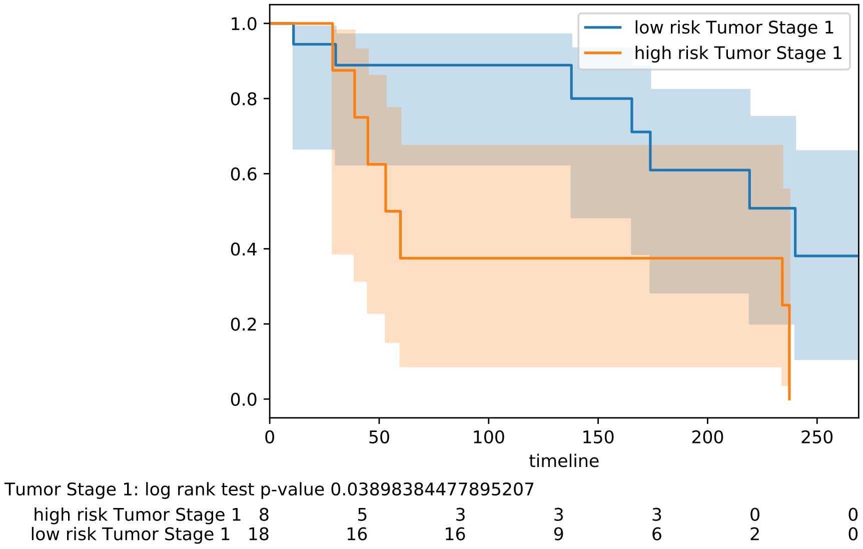

Figure 20: Stratification tumor stage : 1

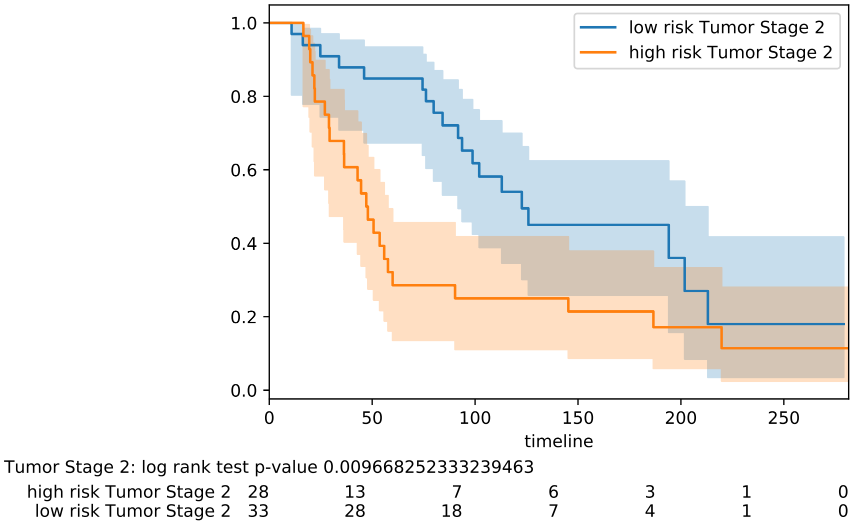

Figure 21: Stratification tumor stage : 2

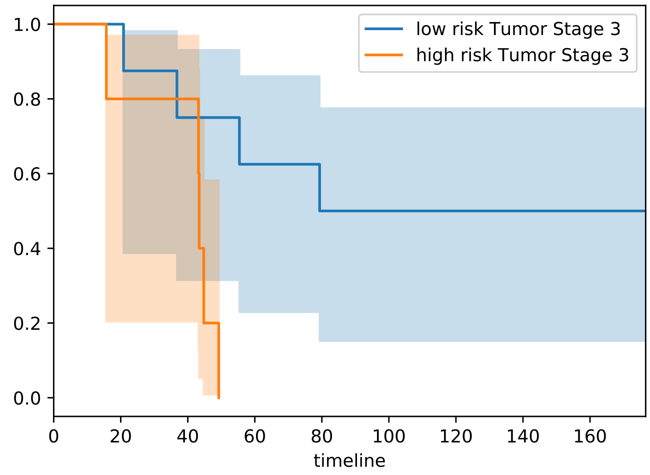

Tumor Stage 3: log rank test p-value 0.02094927141077609

| | | | | | | | | | | |
|---|---|---|---|---|---|---|---|---|---|---|
| high risk Tumor Stage 3 | 5 | 4 | 4 | 0 | 0 | 0 | 0 | 0 | 0 | 0 |
| low risk Tumor Stage 3 | 8 | 8 | 6 | 5 | 4 | 3 | 3 | 2 | 2 | 0 |

Figure 22: Stratification tumor stage : 3

### 2 Overlap of our fingerprints with previously published multi-gene fingerprints

A comparison of our gene panel and the known 33 multigene pannels listed in [1] shows minimal overlap of our fingerprints with previously described fingerprints for breast cancer. We have 1 overlap over 70 genes for Mammaprint fingerprint (Mamma), and 1 overlap for the 16 genes in the Oncotype DX fingerprint (RS).

| Therapy class | gene | Fingerprints |
|---|---|---|
| 'YES', 'NO', 'YES' | G1 | ['MBC'] |
| | G2 | ['p53'] |
| 'NO', 'NO', 'YES' | G3 | ['GGI97', 'Pawitan', 'Robust'] |
| | G4 | ['Novel2'] |
| | G5 | ['GGI97', 'Wang'] |
| 'NO', 'NO', 'NO' | G6 | ['IGS', 'MBC'] |
| | G7 | ['Olaf'] |
| | G8 | ['MBC'] |
| | G9 | ['Mamma'] |
| 'YES', 'NO', 'NO' | G10 | ['MBC'] |
| | G11 | ['HDPP', 'PAM50', 'RS'] |
| 'YES', 'YES', 'NO' | - | - |
| 'YES', 'YES', 'YES' | - | - |
| 'NO', 'YES', 'NO' | G12 | ['Chang'] |
| | G13 | ['LM', 'MBC'] |
| 'NO', 'YES', 'YES' | G14 | ['Wang'] |
| | G15 | ['BCI', 'GGI97', 'PAM50', 'Pawitan', 'Robust'] |

Table 1: Overlap of CVN-based fingerprints and published fingerprints. Fingerprints are indicated with the label used in [1].Common genes are indicated as $G1..G15$.

### 3 Statistics on patient sets

Here we report the distribution of several patients features over training, validation and testing sets (after possible equalization in a therapy class). The distributions are qualitatively similar for the three sets over all the features considered (17 features): intrinsic subtypes, type of breast surgery, histological subtypes, inferred menopausal status, HER2 SNP6 copy number gain/loss, laterality, intrinsic clustering, cohort of origin, ER status by immunohistochemical analysis, age at diagnosis, hormone receptors status, cellularity, lymph node status, tumor stage and tumor grade.

| | Train | | Validation | | Testing | |
|---|---|---|---|---|---|---|
| Num | 326 | - | 186 | - | 200 | - |
| NEG | 113 | 34% | 56 | 30% | 67 | 33% |
| POS | 213 | 65% | 130 | 69% | 133 | 66% |
| no data | 4 | - | 4 | - | 2 | - |

Table 2: Lymphnode status.

| | Train | | Validation | | Testing | |
|---|---|---|---|---|---|---|
| Num | 240 | - | 141 | - | 155 | - |
| 1 | 50 | 20% | 25 | 17% | 27 | 17% |
| 0 | 1 | 0% | 0 | 0% | 0 | 0% |
| 3 | 27 | 11% | 25 | 17% | 25 | 16% |
| 2 | 161 | 67% | 91 | 64% | 100 | 64% |
| 4 | 1 | 0% | 0 | 0% | 3 | 1% |
| no data | 90 | - | 49 | - | 47 | - |

Table 3: Tumor stage.

| | Train | | Validation | | Testing | |
|---|---|---|---|---|---|---|
| Num | 323 | - | 185 | - | 197 | - |
| 1 | 19 | 5% | 5 | 2% | 9 | 4% |
| 3 | 207 | 64% | 120 | 64% | 127 | 64% |
| 2 | 97 | 30% | 60 | 32% | 61 | 30% |
| no data | 7 | - | 5 | - | 5 | - |

Table 4: Tumor grade.

| | Train | | Validation | | Testing | |
|---|---|---|---|---|---|---|
| Num | 328 | - | 187 | - | 202 | - |
| Normal | 27 | 8% | 15 | 8% | 10 | 4% |
| Basal | 75 | 22% | 27 | 14% | 32 | 15% |
| Her2 | 52 | 15% | 27 | 14% | 39 | 19% |
| LumB | 67 | 20% | 50 | 26% | 42 | 20% |
| claudin-low | 37 | 11% | 23 | 12% | 29 | 14% |
| LumA | 70 | 21% | 45 | 24% | 50 | 24% |
| no data | 2 | - | 3 | - | 0 | - |

Table 5: CLAUDIN SUBTYPE

| | Train | | Validation | | Testing | |
|---|---|---|---|---|---|---|
| Num | 326 | - | 186 | - | 200 | - |
| MASTECTOMY | 209 | 64% | 118 | 63% | 130 | 65% |
| BREAST-CONSERVING | 117 | 35% | 68 | 36% | 70 | 35% |
| no data | 4 | - | 4 | - | 2 | - |

Table 6: BREAST SURGERY

| | Train | | Validation | | Testing | |
|---|---|---|---|---|---|---|
| Num | 330 | - | 190 | - | 202 | - |
| IDC+ILC | 11 | 3% | 14 | 7% | 10 | 4% |
| IDC-MUC | 6 | 1% | 6 | 3% | 5 | 2% |
| ILC | 25 | 7% | 9 | 4% | 15 | 7% |
| OTHER-INVASIVE | 1 | 0% | 1 | 0% | 0 | 0% |
| OTHER | 1 | 0% | 0 | 0% | 0 | 0% |
| IDC-MED | 6 | 1% | 2 | 1% | 3 | 1% |
| INVASIVE-TUMOUR | 3 | 0% | 0 | 0% | 1 | 0% |
| IDC-TUB | 6 | 1% | 3 | 1% | 3 | 1% |
| DCIS | 1 | 0% | 0 | 0% | 0 | 0% |
| IDC | 270 | 81% | 155 | 81% | 165 | 81% |
| no data | 0 | - | 0 | - | 0 | - |

Table 7: HISTOLOGICAL SUBTYPE

| | Train | Validation | Testing |
|---|---|---|---|
| num | 330 | 190 | 202 |
| mean | 4.49 | 4.56 | 4.54 |
| std dev | 1.12 | 1.00 | 1.17 |
| median | 4.14 | 5.02 | 5.03 |
| min | 1.03 | 2.01 | 1.05 |
| max | 6.36 | 6.26 | 6.12 |

Table 8: NPI

| | Train | | Validation | | Testing | |
|---|---|---|---|---|---|---|
| Num | 330 | - | 190 | - | 202 | - |
| pre | 111 | 33% | 56 | 29% | 60 | 29% |
| post | 219 | 66% | 134 | 70% | 142 | 70% |
| no data | 0 | - | 0 | - | 0 | - |

Table 9: INFERRED MENOPAUSAL STATE

| | Train | Validation | Testing |
|---|---|---|---|
| num | 330 | 190 | 202 |
| mean | 103.24 | 110.35 | 112.99 |
| std dev | 76.09 | 76.35 | 77.80 |
| median | 88.88 | 102.00 | 98.40 |
| min | 4.17 | 0.10 | 5.83 |
| max | 337.03 | 301.23 | 322.83 |

Table 10: OS MONTHS

| | Train | | Validation | | Testing | |
|---|---|---|---|---|---|---|
| Num | 330 | - | 190 | - | 202 | - |
| NEUT | 224 | 67% | 131 | 68% | 137 | 67% |
| LOSS | 20 | 6% | 8 | 4% | 8 | 3% |
| GAIN | 86 | 26% | 51 | 26% | 57 | 28% |
| no data | 0 | - | 0 | - | 0 | - |

Table 11: HER2 SNP6

| | Train | | Validation | | Testing | |
|---|---|---|---|---|---|---|
| Num | 306 | - | 181 | - | 189 | - |
| r | 140 | 45% | 92 | 50% | 87 | 46% |
| l | 166 | 54% | 89 | 49% | 102 | 53% |
| no data | 24 | - | 9 | - | 13 | - |

Table 12: LATERALITY

| | Train | | Validation | | Testing | |
|---|---|---|---|---|---|---|
| Num | 330 | - | 190 | - | 202 | - |
| 4.5 | 34 | 10% | 19 | 10% | 21 | 10% |
| 10 | 78 | 23% | 36 | 18% | 33 | 16% |
| 1 | 24 | 7% | 17 | 8% | 10 | 4% |
| 3 | 33 | 10% | 22 | 11% | 23 | 11% |
| 2 | 8 | 2% | 12 | 6% | 8 | 3% |
| 5 | 49 | 14% | 27 | 14% | 30 | 14% |
| 4 | 17 | 5% | 5 | 2% | 12 | 5% |
| 7 | 20 | 6% | 11 | 5% | 8 | 3% |
| 6 | 10 | 3% | 7 | 3% | 10 | 4% |
| 9 | 25 | 7% | 16 | 8% | 19 | 9% |
| 8 | 32 | 9% | 18 | 9% | 28 | 13% |
| no data | 0 | - | 0 | - | 0 | - |

Table 13: INTCLUST

| | Train | | Validation | | Testing | |
|---|---|---|---|---|---|---|
| Num | 330 | - | 190 | - | 202 | - |
| 1 | 95 | 28% | 58 | 30% | 57 | 28% |
| 3 | 115 | 34% | 61 | 32% | 73 | 36% |
| 2 | 44 | 13% | 29 | 15% | 36 | 17% |
| 5 | 27 | 8% | 19 | 10% | 14 | 6% |
| 4 | 49 | 14% | 23 | 12% | 22 | 10% |
| no data | 0 | - | 0 | - | 0 | - |

Table 14: COHORT

| | Train | | Validation | | Testing | |
|---|---|---|---|---|---|---|
| Num | 328 | - | 189 | - | 199 | - |
| neg | 150 | 45% | 65 | 34% | 72 | 36% |
| pos | 178 | 54% | 124 | 65% | 127 | 63% |
| no data | 2 | - | 1 | - | 3 | - |

Table 15: ER IHC

| | Train | Validation | Testing |
|---|---|---|---|
| num | 330 | 190 | 202 |
| mean | 56.48 | 58.25 | 57.09 |
| std dev | 13.29 | 14.36 | 12.69 |
| median | 55.30 | 58.97 | 57.38 |
| min | 28.29 | 26.72 | 21.93 |
| max | 90.00 | 96.29 | 84.73 |

Table 16: AGE AT DIAGNOSIS

| | Train | | Validation | | Testing | |
|---|---|---|---|---|---|---|
| Num | 292 | - | 167 | - | 185 | - |
| HER2+ | 53 | 18% | 24 | 14% | 31 | 16% |
| ER-/HER2- | 86 | 29% | 37 | 22% | 43 | 23% |
| ER+/HER2–High-Prolif | 82 | 28% | 65 | 38% | 65 | 35% |
| ER+/HER2–Low-Prolif | 71 | 24% | 41 | 24% | 46 | 24% |
| no data | 38 | - | 23 | - | 17 | - |

Table 17: THREEGENE

| | Train | | Validation | | Testing | |
|---|---|---|---|---|---|---|
| Num | 320 | - | 186 | - | 200 | - |
| high | 180 | 56% | 92 | 49% | 106 | 53% |
| moderate | 108 | 33% | 70 | 37% | 75 | 37% |
| low | 32 | 10% | 24 | 12% | 19 | 9% |
| no data | 10 | - | 4 | - | 2 | - |

Table 18: CELLULARITY

### 4 Features of independent cohorts

Many BC data set are available on the NCBI GEO repository. We have selected 4 data sets using the following criteria. The data set should have a sufficient number of patients (at least 30 patients). The data set should have survival data (preferably Overalls survival) recorded explicitly for each patient. The data set should have explicit adjuvant therapy record for each patient, or the same treatment should be declared common to all patient in the data set. We excluded data set for which the treatment record for each patient could be inferred in principle from other records (e.g. hormonal status) but are not declared explicitly. Data from TCGA (The Cancer Genome Atlas) is not used since the treatment status is known only for few patients, and the TCGA[1] records do not follow the same taxonomy of the Metabric records.

[1]https://www.cancer.gov/about-nci/organization/ccg/research/structural-genomics/tcga

### 5 Measures of performance

*Evaluation of prediction performance.* For a set of patients whose survival data is known, and a prediction has been computed we have a confusion matrix with entries:

1. TP is the number of patients with actual short survival whose prediction is short survival.
2. TN is the number of patients with actual long survival whose prediction is long survival.
3. FP is the number of patients with actual long survival whose prediction is short survival.
4. FN is the number of patients with actual short survival whose prediction is long survival.
5. NR is the number of patients for which the method does not produce a prediction (no response).

Note that patients for which no prediction is given are not counted in any of the four categories standard categories (TP, TN, FP, FN), thus the number of "no reponse" (NR) is a further performance parameter to be considered. For the total number of patients examined $TOT$ it hods:

$$TOT = NR + TP + TN + FP + FN$$

*Slackness (s).*

$$s = \frac{NR}{NR + TP + TN + FP + FN} = \frac{NR}{TOT}$$

The following performance functions are considered:
*(Relative) Accuracy* (Acc)

$$Acc = \frac{TP + TN}{TP + TN + FP + FN}$$

In situations where the number of patients is fixed and the number of "no answers" is within a small range, we can use also use directly the absolute accuracy (TP+TN) as a figure of merit for ranking solutions.
Positive predictive value (PPV)

$$PPV = \frac{TP}{TP + FP}$$

Negative predictive value (NPV)

$$NPV = \frac{TN}{TN + FN}$$

Sensitivity (Sen.)

$$Sen = \frac{TP}{TP + FN}$$

Specificity (Spe.)

$$Spe = \frac{TN}{TN + FP}$$

Odds ratio (OR):

$$OR = \frac{TP \cdot TN}{\max\{1, FP\} \cdot \max\{1, FN\}}$$

Note that this formulation is always well defined even when FP or FN are zero. For OR we also report the 95% confidence interval and the corresponding Fisher's exact test p-value. Cohen's Kappa ($\kappa$).

$$\kappa = \frac{p_o - p_e}{1 - p_e}$$

where $p_o$ is the observed relative accuracy of the method, and $p_e$ is the expected relative accuracy of a random predictor that uses the marginal probabilities. Specifically,

$$p_o = \frac{TP + TN}{TP + TN + FP + FN} = ACC$$

$$p_e = \frac{(TP + FN)(TP + FP) + (TN + FN)(TN + FP)}{(TP + TN + FP + FN)^2}$$

Note that the value is undefined when $p_e = 1$, but this case can can be attained only trivially when the set of patients has only one class of survival. Cohen's Kappa when positive measures the gain in performance of the proposed classifier against the performance of a random classifier using the marginal probabilities, as measured by its expected accuracy. The AUC ROC (Area under the curve of the receiver characteristic curve)[2] measures the average capability of a classifier to discriminate two populations along a scalar value. It can be computed via equivalence to the Wilcoxon Mann Whitney test[3].

[2]https://it.wikipedia.org/wiki/Receiver_operating_characteristic

[3]https://en.wikipedia.org/wiki/Mann_Whitney_U_test

### 6 Method in detail: Construction of Coherent Voting Networks

#### 6.1 Definition of the problem, and objectives

We give her details of the methodology for building, training and using a novel classifier methodology, which we name "Coherent Voting Networks". For concreteness we describe the technique in the context of the prognostic problem for post-surgery breast cancer patients although its description and scope can be cast in more generic terms.
Given a set of $n$ patients $P = \{p_1, ..p_n\}$, and a set of $k$ genes (mRNA) $G = \{g_1, ...g_k\}$, we have at our disposal gene expression measurements for each gene and in each patient[4], organized as a $n \times k$ matrix $M(P, G)$. For each patient we have also the survival function $S : P \rightarrow \{\text{low-risk}, \text{high-risk}\}$ where high-risk indicates survival below five years, and low-risk indicates survival above five years. The classification problem we solve is as follows: use the matrix $M$ and the survival funcrion $S$ to train a classifier, so that for a new patient $p'$ for which gene expression levels for a fingerprint $G' \subset G$ of the genes is known, we can infer the value of the survival function $S(p')$ with high accuracy.
Our objective is to obtain simultaneously (a) a fingerprint $G'$ of small size, and (b) a good performance of the classifier as measured in a train-validation-test evaluation pipeline.[5]

#### 6.2 Coherent voting networks

The new classification method relies on the hypothesis that the input data ($P$,$G$,$M$,$S$) admits a combinatorial construction called a "coherent voting network", such construction is in general not unique, thus among the possible constructions we will perform several optimization and filtering steps for attaining goals (a) and (b).
*Definition of a voting network.* A "voting network" is composed of (I) a collection $\mathcal{C}$ of subsets of $P$, $\mathcal{C} = \{C_1, ...C_h\}$ where $C_i \subset P$ for each $i \in [1, h]$ and (II) a pair $F = (F_1, F_2)$ composed of a first level voting function $F_1$ that maps any restriction of $S$ to a value in $\{\text{low-risk}, \text{high-risk}, \perp\}$, where $\perp$ stands for an undefined label, and a secondary voting function $F_2$, applied to the output of $F_1$.
For a patient $p \in P$, let $\mathcal{C}(p)$ be the sub-collection of sets in $\mathcal{C}$ that contain $p$ (i.e. $\mathcal{C}(p) = \{C \in \mathcal{C} | p \in C\}$). Given $p$, for each set $C \in \mathcal{C}(p)$, the voting function $F_1$ is evaluated on the restriction of $S$ on $C \setminus \{p\}$ (we do not consider $p$ in the input as a patient cannot vote for herself).

[4]Typically via high throughput transcriptomics. For simplicity we assume a full matrix, however imputation of missing values in the matrix is not needed, as missing entries are just missing edges in the graph representation of the matrix.

[5]A train-validation-test, although different from ours, is used as evaluation protocol of the Sage Bionetworks-DREAM Breast Cancer Prognosis Challenge (BCC) conducted between July 2012 and December 2012.

The prediction $Pr(p)$ for $p$ is obtained with the secondary voting function $F_2$, applied to the values of $F_1(C)$ for all $C \in \mathcal{C}(p)$.
The simplest implementation has both $F_1$ and $F_2$ as the majority function (returning $\perp$ in case of tie). However more complex voting schemes will be discussed below.

The voting network $(\mathcal{C}, F)$ is *coherent* for $p$ if $Pr(p) = S(p)$. We have a *fully coherent voting network* when $(\mathcal{C}, F)$ is coherent for each $p \in P$. We have a $\alpha$*-coherent voting network* when $(\mathcal{C}, F)$ is coherent for at least a fraction $\alpha$ of the $|P|$ patients.
The reason why this construction is of interest is that, since the vote for a patient $p$ does not depend on $S(p)$, we can take $Pr(p)$ as the prediction for $p$ when $S(p)$ is not know (or not disclosed to the algorithm). A voting network that is coherent, for a high value of $\alpha$, on its labelled nodes has a good chance of being coherent also for its unlabelled nodes (there will be only one unlabelled node only at any time).

#### 6.3 Construction of a Voting network from the input matrix

The first task is to remove from the matrix $M$ genes that do not have sufficient discriminative power, since for these genes random fluctuations and experimental noise (technical or biological) cover the main signal we wish to exploit.
*Initial statistical filter on the Gene set.* For a gene $g_i$ in $G$ consider the vector $V_i$(low-risk) of entries of $M(g_i, p)$ when $S(p) =$ low-risk, and $V_i$(high-risk) of entries of $M(g_i, p)$ when $S(p) =$ high-risk. We apply to $V_i$(high-risk) and $V_i$(low-risk) a combination of three elementary statistical tests (t-test, Kolmogorov–Smirnov test, and Mann-Whitney U test), and a fold change test, with given fold change and p-value thresholds. Genes passing the tests are retained in the filtered set $G' \subset G$ and a reduced matrix $M(P, G')$ is obtained. This phase requires the setting of a few parameters, including the type of test performed, the maximum p-value for accepting the test, and the threshold for the accepted on the *fold change*.[6]
*Gene expression level quantization.* Gene expression measurements have a large dynamic range of values, and it is unlikely that two patients have the same expression level of a gene, also due to measuring errors. In order to define a discrete structure, we quantize the dynamic range of values of each gene into intervals and we replace each entry in $M$ with the corresponding interval. Intervals are induced by cut-points on the real line. We set a minumum and maximum number of cut points, and a minimum and maximum percentage of patients in each partition induced by the cut-point. We recursively apply cut-point selection methods derived from [4, 5] until the limit of cut points is reached, or some internal stopping criterion is reached. This phase requires the setting of a few parameters, including the type of quantization objective function, the minimum and maximum number of cut points selected, and the minimum and maximum percentage of values in either interval generated by a split point, and the number of significant digits to be considered in the gene expression

[6]Here techniques for Differential Gene Expression analusis could be used, such as ANOVA, or tools like DSeq2 [2], edgeR [3]. However this initial screening phase is in our intention quite light, as subsequent phases should take care of further refining the gene set to attain the final fingerprint.

measurements. Also, such quantization functions need to be able to handle patients for which the survival function is undefined.

*Patient-gene-interval bipartite graph.* After the quantization phase each entry of the matrix $M(P, G)$ is a pair (gene, interval on the real line). We thus build a bipartite graph in which there is a first class of vertices $V_P$, labelled with the patients id. The second class of vertices is $V_{G,I}$ where we have a node labelled with a pair of identifier: the first identifier indicates a gene in $G$, the second identifier is an interval $I$. We set an arc from node $p \in V_P$ to node $(g, i) \in V_{G,I}$ if and only if $M[p, g] = i$. We call this bipartite graph $BG(P, G, I)$.

*Dense Community detection in bipartite graphs.* Similarly to [6], we define a *partial dense cover of radius 2* for the bipartite graph $G(V_1, V_2) = BG(P, G, I)$. A partial dense cover is a collection of dense subgraphs of $G$ with density above a threshold $\delta$ and with a number of nodes of type 1 above a minimum threshold $q_1$ and a number of nodes of type 2 above a minimum threshold $q_2$. We compute this dense cover using a variant of the *Core & peel* algorithm in [6]. The *Core & peel* algorithm is described in [6] for a graph $G$, thus we adapt it to handle properly bipartite graph $B = (V_1, V_2, E \subset V_1 \times V_2)$, by changing accordingly the notion of density, and imposing a that a set in the cover has a minimum number of nodes in class $V_1$ and a minimum number of nodes in class $V_2$. In our application we will have $V_1 = V_P$ and $V_2 = V_{G,I}$ in the bipartite graph $BG(P, G, I)$ Note that each set of vertices in the cover will have both patients and and intervals-gene nodes. Let $\mathcal{D} = \{D_1, .., D_m\}$ be this dense cover. This phase requires the setting of the density threshold (as the percentage of required edges over the maximum possible number of edges on a bipartite complete graph with the same nodes. Typical values range from 0.6 to 0.9. as well as minimum thresholds $q_1$ and $q_2$ (usually set to 3).

*Determination of a voting network from the partial dense cover.* We can obtain from $\mathcal{D}$ a voting network by removing from each $D_i \in \mathcal{D}$ the non-patient nodes (equivalently, projecting the sets in $\mathcal{D}$ on the patient component).

#### 6.4 Sparsification of a partial dense cover

*Community sparsification.* Our objective in this phase of the algorithm is to obtain from $BG$ a bipartite graph $BG'$ for which we have partial dense cover $\mathcal{D}'$ which is similar to $\mathcal{D}$ for the patient component, but uses a much smaller set $G'$ of genes. Thus we obtain from $\mathcal{D}$ a gene set system $GSS$ by removing from each set in $\mathcal{D}$ the patient-nodes and removing the interval component of the remaining nodes. Next we select a covering threshold $c$ and apply a greedy set multi-cover algorithm [7] to $GSS$, this algorithm will produce a set of genes $G'$ with the property that for each $g \in GSS$, either $g \subseteq G'$, or $|g \cap G'| \geq c$. Moreover it has been proved that the number of genes in $|G'|$ is within a logarithmic factor of the smallest subset of $G$ with this property. This phase requires the setting of one main parameter that is the covering number $c$, typically will be a small number chosen between 3 and 7.

*Reduced Patient-gene expression interval bipartite graph.* We now compute a reduced bipartite graph $BG(P, G', I)$ using the previously described procedure on the reduced set $G'$, and we re-apply the community detection procedure to obtain the dense cover $\mathcal{D}'$ and the corresponding voting network. We then test the coherence of the voting network to assess its property of being a $\alpha$-coherent voting network (for one of the voting functions). If this is verified this voting network can be used for classification purposes.

*How to turn a $\alpha$-coherent voting network into a classifier.* Intuitively, we assume that the algorithm $A$ that produces $(\mathcal{C}, F)$ starting from $(P,G,M,S)$ and a fixed vector of parameters, is sufficiently stable under small perturbations of the input. We apply the same construction to $(P \cup \{p\},\ G',\ M(P \cup \{p\}, G'),\ S \cup \{p \to \bot\})$. When we control the coherence of the resulting output $(\mathcal{C}', F)$, the level of coherence $\alpha$' can be computed for all patients in $P$. We check that $\alpha' \approx \alpha$ in order to accept $Pr(p)$ as the prediction for the new patient $p$, based on the coherence of the network on the primitive set of patients.

### 7 Extending the repertoire of voting schemes

The primary voting function $F_1$ is chosen among one of the following standard voting functions:

a) "unanimity": the voting function returns label $X \in \{\text{low-risk}, \text{high-risk}\}$ if there is at leat one patient with label $X$ and all patient for which the label is not undefined have the same label $X$.

b) "qualified majority at threshold $t$:

c) "simple majority": the voting function returns label $X \in \{\text{low-risk}, \text{high-risk}\}$ if there is at leat one patient with label $X$ and the number of patients with the same label $X$ is larger than for the number of patients with any other defined label.

d) "more than expected": the voting function returns label $X \in \{\text{low-risk}, \text{high-risk}\}$ if the number of patients with label $X$ is larger than the expected number of patients with label $X$ in a random sample of patients from $P$, of the same size, with p-value $p \leq 0.05$. This voting function is important when the Training data has unbalanced classes.

e) given the survival function $S : P' \to \{\text{low-risk}, \text{high-risk}\}$ we compute a weighting function on the elements of $\{\text{low-risk}, \text{high-risk}\}$ by counting the number of patients with a given label. Formally the wight of label $X$ is $|\{p \in P' | S(p) = X\}|$

The secondary voting function $F_2$ is chosen among:

f) "simple majority", when the primary voting function is one of a), b), c), d).

g) "weighted majority": when the primary voting function is e), we sum the weights for each label, and return the label with highest cumulative weight.

h) "unanimity plus weighted majority": We apply the simple majority on the primary vote done with the "unanimity" function, if the result is defined we return it, otherwise we apply the "weighted majority" and we return this value.

This phase requires the setting of one main parameter that is the voting policy.

### 8 Method in detail: Training and optimization of Coherent Voting Networks

*Train-validation-test evaluation pipeline.* The cohort METABTRIC of roughly 2000 patients is split randomly into three sets with proportions (1/2, 1/4, 1/4): training set (roughly 1000 patients), validating set (roughly 500 patients, and testing set (roughly 500 patients). We remove patients for which the survival class cannot be determined. We further stratify each set of patients by the available therapy information. The adjuvant therapy information is whether the patient has received endocrine therapy (YES/NO), radiation therapy (YES/NO) or chemotherapy (YES/NO). This stratification produces eight subclasses of patients. In each subclass of patients we check the ratio of the size of the two survival classes $\{\text{high-risk}, \text{low-risk}\}$. When the ratio of the largest to the smallest class was greater than 2.5, the larger class was sub-sampled so to re-balance it within the 2.5 ratio.
The train-validation-pipeline is applied for each class separately on the corresponding three sets of patients. Let us denote the tree sets of patients for a class as $Train$, $Val$, and $Test$. Our aim is to find a small fingerprint, to optimize the CVN hyperparameters, and to evaluate the performance of the chose classifier. A configuration $C$ is a pair $(G', Par)$ where $G'$ is a set of genes (as obtained by the sparsification procedure) and $Par$ is a vector of parameters that are given as part of the input to the several phases of the algorithm. Each field of $Par$ takes values in a domain of small discrete size. The vector space of all possible parameters is denoted by $PAR$.
*Generation of fingerprints.* For a given fixed vectors of parameters $Par \in PAR$ we obtain from the input ($Train$, $G$, $M(Train, G), S(Train)$ a voting network and a panel of genes $G'$ after sparsification. Repeating this operation for all the vectors of parameters in vector space of parameters ($PAR$) we obtain a pool of candidate gene panels $G'$ (and corresponding vector of parameters and voting networks).
*Post training selection of fingerprints.* We aim at reducing the number of candidate gene panels by retaining only a few high quality candidate gene panels. We discard a candidate panel $G'$ if it is too large (we apply a cutoff value of 20 genes). We discard a candidate panel $G'$ if the quality parameter ($\alpha$) is below a factor 0.85 of the top value for $\alpha$ among all the generated voting networks. We sort the so filtered fingerprints by their value of $\alpha$ for the associated voting network and we take at most the top ranking 30 fingerprints, after removing eventual duplicate candidate panels.
*Validation and Testing.* In this phase we aim at finding a configuration (fingerprint and parameters) the balances the performance on the LOOCV (leave-one-out-cross-validation)

measures for the Train set, on the train-test evaluations of the Validation data , and on the train-test evaluations of the Test data, with a minimal usage of Test data (as quantified by the look ahead number).
A straightforward sorting by just one of the quality measure does not give the desired outcome, therefore we use a more complex schema that uses in intermediate steps Pareto frontiers[7] and Pareto stratifications of the (multi dimensional) performance data vectors, and lower bound filters to discard outliers. For this part of the method by *score* we intend either the kappa statistic or the OR statistic. A configuration is represented by a point in 3D space corresponding to the coordinates (accuracy, score, antislack).[8]

a) Automatic lower bound computations. Based on Validation data only we compute Pareto strata (iterated the Pareto fronts) up to collecting at least 8 unique points we compute for these points the minimum number of hits (accuracy) and the minimum score, subject to slack below 0.2. These two values for accuracy and score form the filtering values are computed on all the Validation data.

b) For each candidate fingerprint separately, we retain the points (accuracy, score, antislack) on Validation data that satisfy the lower bounds, and we compute the principal Pareto frontier of this subset of points.

c) We collect all such Pareto frontiers in a single set, removing duplicate points (i.e. if two configuration correspond to the the same point only one is retained).

d) This reduced set is then re-split into Pareto strata but the point associated to a configuration is now based on the LOOCV of Train data of the corresponding configurations, and each stratum is sorted by the LOOCV score value. In a variant of this phase the points may be constructed as coordinate-wise arithmetic mean (or geometric mean) of the LOOCV of Train points, and the corresponding points for the Validation data.

e) In the order of the strata and the internal order of each stratum, we check the stopping criterion. The numbers of these checks is to *look ahead number* since the stoping criterion involves both Validation and Test data.

f) We stop the look ahead procedure if the Test score value is larger than the Validation score value, or if Test score value is smaller but within a relative displacement of less than 0.2 form the Validation score value.

### 9 Manual fingerprint and hyperparameter optimization

For some of the therapy classes the *Validation and Testing* method outlined above fails to produce a significant result, either because the lower bounding procedure a) is too aggressive,

[7]https://en.wikipedia.org/wiki/Multi-objective_optimization
[8]Antislack is 1 minus the slack factor.

or the stopping criterion in f) does not succeed. For these cases we resort to a manual selection, by choosing via visual inspection a pool of high performance configurations on the Validation data. We then *regularize* this set of configuration by taking the Cartesian product of the projections of the configurations on its components. This new set of configurations is then checked on the Test data, and the best performing result on test data is reported. Note that in this case the lookahead number is relatively large, in the order of a few tens. However such manually selected fingerprints are of interest since they did perform well on some of the independent set in LOOCV tests.

### 10 Avoiding overfitting

When using minimization of cross validation error in order to perform hyper-parameter optimization (hypothesis selection), we may incur in a form of "overfitting" due to the fact that for a large number of hypothesis and in the presence of noisy data, the minimum may be attained by fitting the noise in the data, rather than the underlying signal [8]. This phenomenon is discusses by A. Y. Ng [8] who also proposes a principled way of choosing an alternative (non minimal) hypothesis from the cross validation that has more chances of a better generalization error. Here we do not implement the method by A. Y. Ng, but we observe that in our specific problem an hypothesis that balances well the cross validation error and the generalization error is found very close to the top of the ranking we build, so that it can be identified with just a few lookahead operations.

### 11 Data, Software and tools used

The code is written in python 2.7. Kaplan–Meier Plots and log rank tests are produced with the python package *lifelines* (lifelines.readthedocs.io/en/latest/). AUC and Odds ratio are computed with the python package *scipy.stats* (www.scipy.org/). Autoweka code has been downloaded from github.com/automl/autoweka.
Data pre processing and post processing has been done on a Intel Core i7 (2.8 GHz) processor with 6GB RAM and 240 GB HD with Windows 10 Pro (64 bits). Generation and optimization of CVN has been run on Linux server with 8 Intel Xeon E5-4620 v2 @ 2.60GHz (total 64 cores) and 500 Gb RAM with CentOS Linux 7 (Core).
Metabric BC data has been downloaded from cBioPortal (www.cbioportal.org). Data on independent cohorts has been downloaded form the NCBI-GEO portal (www.ncbi.nlm.nih.gov/geo/).
Input data files used in train-validate-testing are available at:
https://github.com/MarcoPellegriniCNR/Coherent-Voting-Network-for-BC-prognosis.
